# Using a crowdsourcing open call, hackathon and a modified Delphi method to develop a consensus statement and sexual health survey instrument

**DOI:** 10.1101/2020.10.02.20205542

**Authors:** Eneyi E. Kpokiri, Dan Wu, Megan L. Srinivas, Juliana Anderson, Lale Say, Osmo Kontula, Noor Ani Ahmad, Chelsea Morroni, Chimaraoke Izugbara, Richard de Visser, Georgina Yaa-Oduro, Evelyn Gitau, Alice Welbourn, Michele Andrasik, Wendy V. Norman, Soazig Clifton, Amanda Gabster, Amanda Gesselman, Chantal Smith, Nicole Prause, Adesola Olumide, Jennifer T. Erausquin, Peter Muriuki, Ariane van der Straten, Martha Nicholson, Kathryn A. O’Connell, Meggie Mwoka, Nathalie Bajos, Catherine H Mercer, Lianne Marie Gonsalves, Joseph D. Tucker

**Affiliations:** Faculty of Infectious and Tropical Diseases, London School of Hygiene and Tropical Medicine, London, UK; Institute of Global Health and Infectious Diseases, University of North Carolina, Chapel Hill, NC; Department of Anthropology, School of Global Health, University of North Carolina, Chapel Hill, NC; Department of Sexual and Reproductive Health and Research including the UNDP/UNFPA/UNICEF/WHO/World Bank Special Programme of Research, Development and Research Training in Human Reproduction (HRP) World Health Organisation, Geneva, Switzerland; Population Research Institute, Family Federation of Finland, Helsinki, Finland; Institute for Public Health, Ministry of Health Malaysia, Setia Alam, Shah Alam, Selangor, Malaysia; Botswana Sexual and Reproductive Health Research Initiative, Botswana Harvard Partnership, Garborone, Botswana; Department of International Public Health, Liverpool School of Tropical Medicine, Liverpool, United Kingdom; International Center for Research on Women (ICRW), Washington DC, USA; School of Psychology, University of Sussex, Brighton, United Kingdom; Centre for Gender Research, Advocacy and Documentation, Department of Sociology and Anthropology, University of Cape Coast, Cape Coast, Ghana; African Population and Health Research Center, Nairobi, Kenya; Salamander Trust, London, United Kingdom; Vaccine and Infectious Disease Division, Fred Hutchinson Cancer Research Center, Seattle, United States; Department of Global Health, University of Washington, Seattle, United States; Department of Family Practice, Faculty of Medicine, University of British Columbia, Vancouver, Canada; Faculty of Public Health & Policy, London School of Hygiene & Tropical Medicine, London, United Kingdom; Centre for Population Research in Sexual Health and HIV, Institute for Global Health, University College London, London, United Kingdom; NatCen Social Research, London, United Kingdom; Gorgas Memorial Institute for Health Studies, Panama City, Panama; The Kinsey Institute, Indiana University, Bloomington, United States; Maternal, Adolescent and Child Health Institute, Durban, South Africa; Liberos, Los Angeles, United States; Institute of Child Health, College of Medicine, University College Hospital, Ibadan Nigeria; Department of Public Health Education, University of North Carolina, Greensboro, NC, United States; Women’s Global Health Imperative, RTI International, Berkeley, CA, United States; Center for AIDS Prevention Studies, Department of Medicine, University of California, San Francisco, United States; Marie Stopes International, London, United Kingdom; EngenderHealth, Program Impact, Research and Evaluation, Washington DC, United States; INSERM (Institut National de la Santé et de la Recherche Medicale), CESP Centre for Research in Epidemiology and Population Health, Gender, Sexual and Reproductive Health Team, Kremlin Bicetre, Paris, France

**Keywords:** sexual health, survey instrument, population representative, global, crowdsourcing

## Abstract

Population health surveys are rarely comprehensive in addressing sexual health, and population-representative surveys often lack standardized measures for collecting comparable data across countries. We present a sexual health survey instrument and implementation considerations for population-level sexual health research. The brief, comprehensive sexual health survey and consensus statement was developed via a multi-step process (an open call, a hackathon, and a modified Delphi process). The survey items, domains, entire instruments, and implementation considerations to develop a sexual health survey were solicited via a global crowdsourcing open call. The open call received 175 contributions from 49 countries. Following review of submissions from the open call, 18 finalists and eight facilitators with expertise in sexual health research, especially in low and middle-income countries (LMICs), were invited to a 3-day hackathon to harmonize a survey instrument. Consensus was achieved through an iterative, modified Delphi process that included three rounds of online surveys. The entire process resulted in a 19-item consensus statement and a 10-minute sexual health survey instrument. This is the first global consensus on a sexual and reproductive health survey instrument that can be used to generate cross-national comparative data in both high-income and LMICs. The inclusive process identified priority domains for improvement and can inform the design of sexual and reproductive health programs and contextually relevant data for comparable research across countries.

**Key points:**

– National population-representative surveys assessing sexual practices, behaviours and health-related outcomes focus on high-income countries and different sexual health measures are often used.
– There is a lack of population-level representative sexual health research in low- and middle-income countries.
– Existing comparable data on sexual practices and behaviours across countries are limited due to the absence of a standardized global sexual health survey instrument.
– We report the global consensus on a set of core sexual health items within a 10-minute survey instrument that accommodates the needs and priorities of people from LMICs and various legal and cultural contexts across countries.
– The consensus process breaks new ground in terms of incorporating feedback from diverse individuals using a crowdsourcing open call and hackathon process.

---

Sexual health is an integral part of overall health and well-being.^1,2^ Understanding sexual practices and behaviours are necessary to design appropriate services for populations and to monitor the impact of interventions. Comparable, cross-national, population-representative data can help to address social determinants of health,^3-5^ better understand social norms related to gender and sexuality,^6^ and improve sexual health systems. However, such data on sexual health are limited.

Many national population-representative surveys assessing sexual practices, behaviours and health-related outcomes focus on high-income countries (HICs).^7-13^ These surveys often use different sexual health measures, making cross-national comparison difficult. In low and middle-income countries (LMICs), some key indicators are captured in standardized national surveys, such as the Demographic and Health Surveys (DHS) and Multiple Indicator Cluster Surveys (MICS).^14,15^ However, these instruments go beyond sexual behaviours and collect few indicators on sexuality.^16^ Additionally, most existing survey instruments were created by experts from HICs with limited feedback from LMIC researchers or communities. Certain sub-groups are particularly under-represented, such as women, sexual minorities, and people with disabilities.^17-21^ Also, social acceptance and cultural beliefs towards sexual health and practices vary by geographical regions and social groups. Thus, priorities of key domains for a sexual health survey differ greatly across countries. Furthermore, access to means of data collection vary, making administration of long instruments especially difficult in some LMIC settings. These issues indicate a need for global expert consultation to seek a consensus on what measures should be included in a global sexual health instrument and guidance on its implementation.

## Methods

Three key methods were employed including a crowdsourcing open call for ideas, a hackathon, and an iterative modified Delphi exercise (Figure 1). Crowdsourcing open calls invite individual participants or groups with a wide range of backgrounds to offer a solution, identify solutions, and share with the wider community.^22,23^ The purpose of the crowdsourcing open call was to solicit existing survey instruments and measures assessing sexual practices, behaviours and health-related outcomes, as well as implementation considerations. A hackathon or designathon is a sprint-like event that brings together individuals with diverse backgrounds to solve a problem.^24^ A hackathon can tap into participants’ experiences and expertise to generate high quality outputs in a transparent and systematic way.^25^ The purpose of this sexual health hackathon was to harmonise exceptional entries received during the open call and deliberate on key items to be included in the survey, aiming to assemble a draft 10-minute sexual health survey at the end of the hackathon. The Delphi method is an iterative multi-stage process used to achieve expert consensus on a subject.^26^ The purpose of this method was to develop consensus statements on the design, training and implementation of a sexual health survey, and to finalize items to be included in the sexual health survey instrument. Each of these methods provided an opportunity for participant engagement to enhance collaboration. This manuscript documents this process and presents the resulting draft survey instrument and consensus statements.

**Figure 1:**
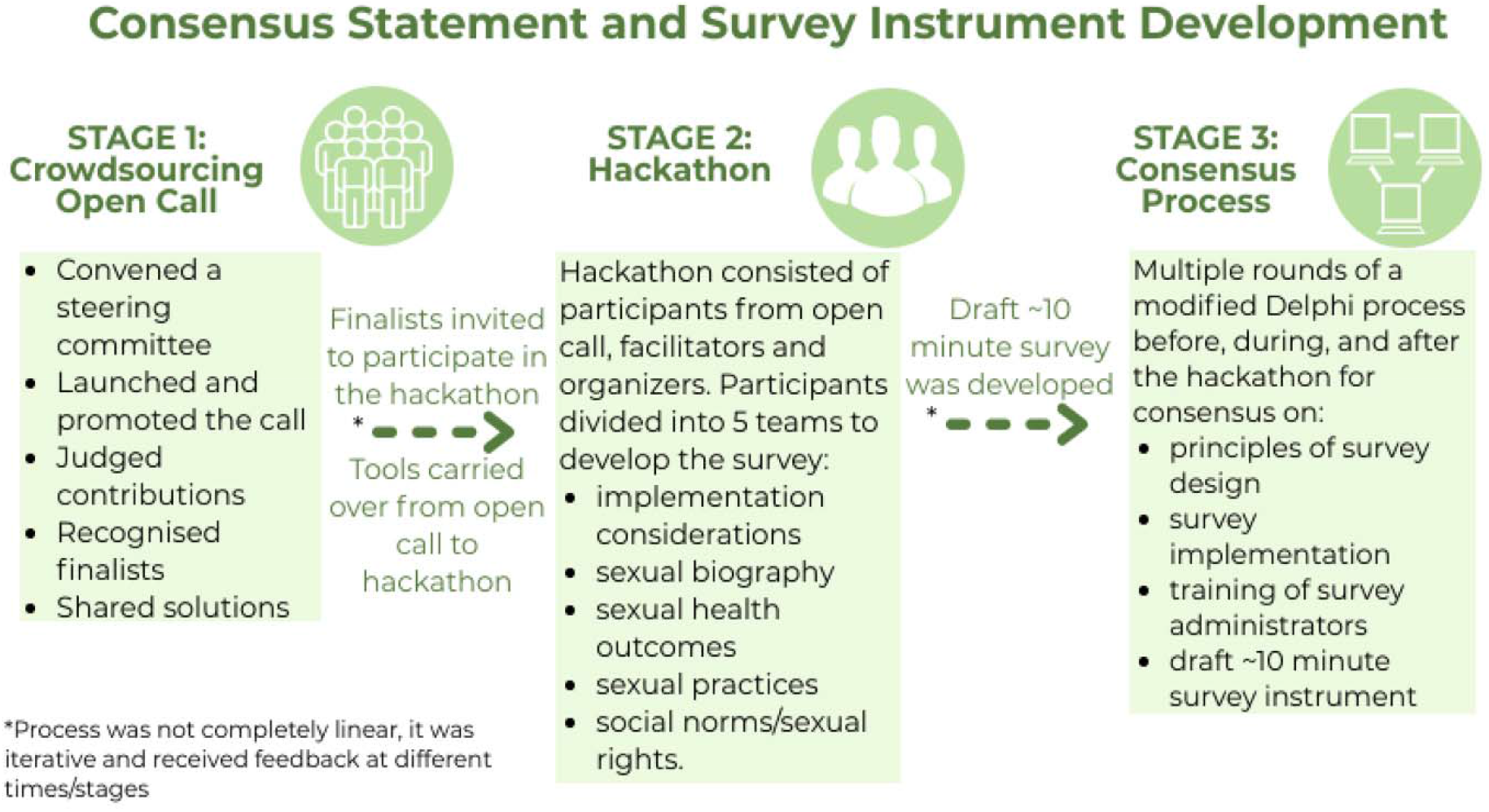
Key components of the consensus process.

## Results

### Crowdsourcing open call

For this call, the community of interest was the diverse community of researchers, leaders, program implementers, and care providers who work in sexual and reproductive health (including family planning, and STI prevention communities) as well as HIV prevention, control and care. The call for ideas (Supplementary file 1) was launched on September 4, 2019 and remained open until November 1, 2019. It was hosted on the WHO/HRP official website and was promoted by partner organisations, including at a special symposium at the 24^th^ Congress of the World Association for Sexual Health in October 2019. The call was translated into Spanish and we accepted contributions in all six official languages of the WHO (Arabic, Chinese, English, French, Russian and Spanish).

At the end of the call, all contributions were screened for eligibility and judged using pre-specified criteria (Supplementary file 2). The HRP open call received 175 total submissions from 49 countries, of which 59 submissions were received from LMICs. Participants came from all six WHO regions including the Americas (85), Europe (38), Africa (25), Eastern Mediterranean (10), South-East Asia (10) and Western Pacific (8). We received six entries in Spanish and two entries in French, all of which were translated into English for screening and judging. After initial screening, 139 unique entries were eligible for judging. Twelve independent judges reviewed submissions. Each submission was reviewed by at least four judges and numerically scored on a 1-10 scale, 10 being the best. Scores for each contribution were averaged and those with a standard deviation greater than 2.5 were reviewed by two additional judges. After collating judge scores, 47 entries achieved a mean score of 7 or greater, emerging as semi-finalists. These were further reviewed by the steering committee who ultimately selected18 finalists based on the mean score achieved coupled with the desire to balance participant demographics and experience working in HIC and LMIC settings. Among finalists, 83% (15/18) had LMIC sexual health research experience. This group included principal investigators on LMIC sexual and reproductive health studies, data analysis experts, sociologists, demographers, epidemiologists, reproductive health leaders, and others with experience in developing national surveys and analysing multi-country data. Finalists were then invited to attend the following hackathon in January 2020.

### Hackathon

This hackathon was jointly organised by the team members at the London School of Hygiene and Tropical Medicine (LSHTM), WHO/HRP, and hosted by the African Population and Health Research Center (APHRC) in Nairobi, Kenya. Other hackathon participants were organizers from WHO/HRP, London School of Hygiene and Tropical Medicine, French National Institute for Health and Medical Research and the host APHRC. In total, 35 individuals participated in the hackathon (Table 1). Participants included 7 organizers from the partner organisations, 10 facilitators, and 18 finalists from the open call. Facilitators were more senior sexual health researchers and experts with extensive research in developing and implementing large population representative surveys such as Demographic Health Surveys (DHS),^14^ the British Natsal,^10,11^ the French ACSF,^12^ and Finnish FINSEX.^13^ Participants were provided with documents to review prior to the hackathon, including themes analysed from contributions to the open call, other relevant sexual health surveys, and a hackathon guide (Supplementary file 3). The hackathon event ran for three days (January 14-16, 2020), with detailed agenda and expected outcomes presented in the hackathon guide. Participants were divided into five small groups of five or six members. Group topics included survey implementation considerations, sexual biography, sexual health outcomes, sexual practices, and social norms/sexual rights. Each group had one facilitator, one organizer, and three or four finalists from the open call. Two additional lead facilitators rotated across all five groups and helped to provide guidance and resolve conflicts arising during group discussions. Each group was given additional parameters for inclusion (see Table 1). Groups were asked to prioritize items for a 10-minute survey and to propose measures already used and standarzied in previous surveys. Groups presented their sections at the end of each day for feedback and discussion.

**Table 1.**
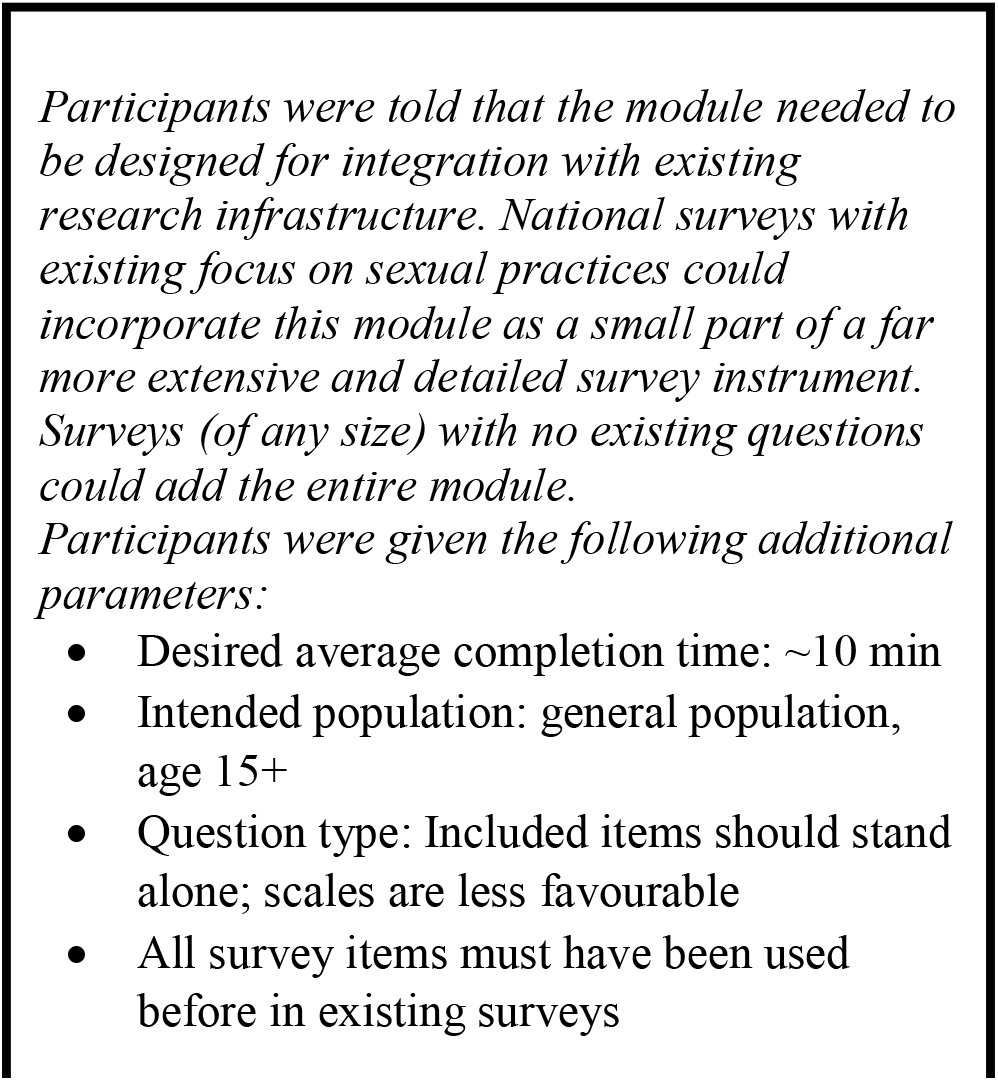
Survey module design.

### Modified Delphi

A multi-round modified Delphi was also completed, with each round informing the next (Supplementary file 4). A five-point Likert scale from strongly agree to strongly disagree was used to record responses. The definition of consensus was set at ≥80% achieved for agree and/or strongly agree. The first round of consensus building focused on establishing statements on the principles for survey design, survey implementation, and training of survey administrators. These were identified and extrapolated from open call submissions. These statements were intended to guide and inform sexual health researchers and implementers towards standardized procedures when conducting sexual health surveys. The first round was conducted just before the hackathon event and included all participants of the hackathon and volunteers identified through the open call. The results from the first round of the consensus statement survey was provided to participants at the hackathon. Statements were revised based on feedback from the first round of the survey. The second round of the consensus statement survey was undertaken during the hackathon event and included both statement items and potential sexual health survey items. This second round was completed by hackathon participants only. The third and final round of the consensus statement and sexual health items survey was conducted after the hackathon via email correspondence and it included the revised consensus statements and the draft items selected for the sexual health survey during the hackathon. Participants invited to provide feedback in this round included all participants and facilitators in the hackathon, members of the steering committee and participants on the open call with a mean score greater than or equal to five. For the consensus statement, participants graded each of the statements. Items that achieved 100% agreement were graded as “U” (unanimous), 90-99% agreement were “A”, and 80-89% agreement were “B”, and items with less than 80% agreement were not included. The steering committee reviewed all grading and made final decisions.

Sixty people were invited to take part in the first-round online survey focused on consensus statements and 47 (78%) responded. This survey included 12 statements on principles of sexual health survey design (7), training (2) and implementation (3). Participants who responded indicated expertise in survey design, piloting, data management, data analysis and field work. Two statements on the design stage did not reach 80% agreement and were revised for the second round. The second round of the survey, focusing on consensus statements and draft sexual health items, was conducted at the start of the hackathon and included 31 participants, with a 100% response rate. Of these participants, 22/31 (71%) had LMIC sexual health research experience.

The final round of the survey included 19 consensus statements (Table 3) alongside the draft sexual health survey instrument. A total of 35 people were contacted and 23 responded with a 66% response rate. All items on the consensus statements achieved ≥80% agreement, and 66/71 items on the survey instrument achieved 80% agreement. Items with lower agreement levels were presented and discussed with the steering committee to either remove or revise. The resulting draft sexual health items survey instrument is included as Supplementary File 5 and the consensus statements are provided in Table 3.

**Table 2:** Characteristics of the hackathon participants.

| Characteristics | Number (n=35) |
| --- | --- |
| **Participant's sex** |  |
| Male | 7 |
| Female | 28 |
| **Role in sexual health research** |  |
| Survey leadership | 19 |
| Survey design | 26 |
| Survey piloting | 23 |
| Data analysis | 28 |
| Administration | 29 |
| **Years of sexual health experience** |  |
| 1-5 | 5 |
| 6-10 | 7 |
| 11-20 | 10 |
| >20 | 13 |
| **Field research experience** |  |
| LMICs | 14 |
| HICs | 13 |
| LMICs and HICs | 8 |
| **LMICs: Low- and middle-income countries; HICs: High income countries** |  |

**Table 3.** Consensus Statements (19 items)

| **Table 3. Consensus Statements (19 items)** |  |  |
| --- | --- | --- |
| **Number** | **Statement** | **Grade** |
| *General principles that apply to design, implementation (including identifying and training interviewers), and dissemination* |  |  |
| A sexual health survey instrument should do the following: |  |  |
| **1.** | Draw on a holistic view of sexual health, as described by the WHO's working definition. | **U** |
| **2.** | Recognize the potentially sensitive nature of certain concepts and be informed about local and national norms and laws related to age of | **U** |
|  | consent, same-sex relationships, abortion, sexual violence, gender issues, and related macro-level factors. |  |
| **3.** | Engage local multi-sectoral key stakeholders across all stages of the survey research project including design, implementation, and dissemination. Key stakeholders might include potential research participants, government officials from across the socioeconomic and political spectrums, policymakers, members of civil society, and others depending on the context. | **U** |
| **4.** | Ensure the survey and its data are used in ways that promote, protect, and fulfil human rights, including sexual rights, per the WHO's working definition (here). | **U** |
| **5.** | Be adaptable to the local population's priorities, needs, norms, and practices. | **U** |
| *Design stage* |  |  |
| **6.** | Capture information on one's sexual and reproductive health, related choices, and outcomes. | **U** |
| **7.** | Reflect the lived reality of the participant taking part in the survey in their local context. | **A** |
| **8.** | Acknowledge the broader determinants of sexual and reproductive health outcomes per the WHO's working definition (here). | **U** |
| **9.** | Include young people under age 18 if in line with local regulations, laws, and ethical norms. This may benefit from discussions with the local ethical review committee whose approval would be required prior to starting research. | **A** |
| **10.** | Avoid language that is derogatory or discriminatory as informed by the local community; use people-centered language (e.g., 'people with disabilities' instead of 'disabled people'). | **U** |
| *Implementation (Identify and Train Interviewers)* |  |  |
| **11.** | Select interviewers who understand the local context. Special consideration should be given to including interviewers with knowledge of or experience with subgroups of participants identified as important by the research team (e.g., older people, sexual minorities, people with physical or mental disabilities, etc.). | **U** |
| **12.** | Core topics of interviewer training include protecting participants, rapport building, the socio-legal environment, ethics training, gender dynamics (e.g., women interviewing men or vice-versa), age dynamics (e.g., younger people interviewing older people), trauma-informed care, and quality control. | **U** |
| **13.** | Core competencies of interviewers include asking obtaining participant consent/assent (for minors), sensitive questions, understanding behaviours considered illegal, managing participant responses to sensitive issues, avoiding biasing participant responses, and demonstrating a non-judgmental demeanor. | **U** |
| **14.** | Training should focus on building mutual understanding between the participant and the interviewer, using participatory training methods where appropriate (e.g. role-playing and/or implicit bias training). There should be regular ongoing supervision and support for interviewers in order to address issues that arise during data collection, particularly when asking about sensitive issues, such as sexual abuse or gender violence. | **A** |
| **15.** | Interviewers must be trained in their legal duties regarding reporting requirements (e.g. with regards to sexual violence, consensual sexual activity among adolescents, even parental consent to access sexual and reproductive health referral services) and ethical duties. The research team should be aware that their actions or omissions may carry legal implications. If a conflict arises between a legal obligation and an ethical duty, the research team should obtain advice from their professional association on how best to proceed and, ultimately, choose to always act in an ethical manner. When relevant issues are identified, the research team must provide information on appropriate services and assist in linking those affected to these services (e.g., legal services, local hotlines, shelters, health and social services) and consider the safety of those affected when dealing with mandatory reporting requirements.<sup>1</sup> | **U** |
| **16.** | Ensure the confidentiality and privacy of participants. | **U** |
| *Dissemination* |  |  |
| **17.** | Create a summary of the research findings accessible to participants. | **U** |
| **18.** | Create a summary of research findings to be shared with policy-makers, public audiences, or others. | **U** |
| **19.** | Work in partnership with local communities to disseminate research findings to key stakeholders as defined above. | **U** |
| U (Unanimous)=100%; A= 90-99% agreement |  |  |

<sup>1</sup>Researchers may be legally required to report certain types of violence or sexual activity to relevant authorities, even though this reporting may conflict with the ethical obligation to protect participants' confidentiality and respect their autonomy (see “Special considerations related to mandatory reporting requirements”, below). It is essential that researchers understand and plan appropriately for situations in which mandatory reporting requirements may apply, recognizing that different standards apply across countries. They will need to explain the limits of confidentiality to research participants. In addition, it may be ethically appropriate to screen participants for immediate safety concerns and to refer them directly to additional support services for their own and their children's safety and well-being.

## Discussion

The global sexual health survey instrument along with a consensus statement and implementation considerations is intended for use in diverse global settings to facilitate cross-country comparisons. It provides a set of core sexual health items resulting in a 10-minute survey instrument and implementation guidance that can be flexibly adapted according to local cultures and contexts. The global consensus was reached by a combination of engagement strategies. These engagement activities empowered and involved sexual health experts from many research fields and backgrounds, especially LMIC experts. We believe this survey would be relevant in various legal and cultural contexts across countries.

We achieved high agreement levels regarding principles for the design process of a national sexual health survey, local capacity building and training of organizers, and implementation principles. Some items related to sensitive issues (e.g., types of sexual behaviours, including same sex behaviours, and sexual violence) will need to be field tested in local settings to understand how best to implement.

Our process underlined the need for further research and measures development for social norms related to sex and sexuality and sexual rights. A wide range of aspects related to social norms were discussed and we narrowed down to eight sub-domains (Supplementary file 5) that were considered important topics shared across different contexts. These eight sub-domains were social beliefs of sex education, contraception, abortion, sexual needs, and same-sex relationships as well as gender norms surrounding consent to sex, premarital sex, and sexual pleasure. Reaching consensus on these indicators for measuring social norms and sexual rights was particularly challenging compared to other domains. We determined two main barriers. First, many important social norm constructs were measured using scales too lengthy for this brief instrument, including the Sexual Consent Scale,^27^ Gender Equitable Men scale,^28^ and Intimate Partner Violence Attitude Scale.^29^ Hence, our brief survey excluded many survey items simply because of length, not because the topic was unimportant. Further research on devising and validating short-version scales to measure these indicators is needed. Second, these sub-themes are strongly associated with local beliefs and cultures, and priority themes are contextually relevant. This highlights the need for cognitive testing and further comparative research.

Moving forward, we encourage the broader research community to comment on the survey via an open review process in which the instrument will be placed on the WHO/HRP website. Experienced, in-country researchers from around the world will be invited to conduct cognitive testing on the instrument. We recommend researchers to include a local group of participants with diverse socio-demographic backgrounds (e.g., gender, age, education, sexual orientation) in cognitive interviews to obtain feedback on survey content and flow, comprehensibility, wording, cultural appropriateness, and length. After amendments to the survey instrument are made based on the interviews, a representative survey can follow to estimate completion time and optimal sample sizes in local contexts.

Our process has some limitations. A wider engagement of audiences from some subgroups (e.g., low-income countries in Asia) to the open call could have led to more submissions from these nations. However, we had strong representation of people undertaking LMIC research across all regions. The open call and hackathon were organized using the English language. However, we invited submissions from all official WHO languages and had hackathon participants fluent in Spanish and French review the respective survey instruments. Third, our process involved an in-person hackathon event which would be more difficult in the COVID-19 era. At the same time, many hackathons have transitioned to digital formats to organized COVID-19 responses, suggesting an alternate pathway.^30^ This suggests that digital hackathons may be able to accomplish the same goals without the risk of in-person activities.

Other strengths of our process included the wide and iterative engagement from a range of professional disciplines related to sexual and reproductive health in a range of cultural settings, the involvement and commitment of leading national and international health organizations, and the strong consensus achieved on quality items throughout the phases of development. This standardized instrument and consensus statement has implications for policy, practice, and research. The instrument can help inform local policy makers and SRH researchers about priority domains for improvement in the local context. Then, it can be used to collect data on sexual and reproductive health-related norms and practices at the population level in order to guide stakeholders to design and implement responsive services and programs to improve SRH. The crowdsourcing approach that we used to develop this survey instrument contrasts conventional guideline development and could lay the foundation for a more participatory consensus statement development process. Research comparing the crowdsourcing approach to conventional approaches is needed.

### Conclusion

We successfully recruited a wide range of experts to engage in rigorous, tested participatory approaches. We achieved consensus on a 10-minute module for a global sexual health survey instrument and on guiding implementation strategies. Our sexual health survey instrument could provide comparable indicators across settings, and has implications for policy, practice, and research. Our survey instrument could also allow flexibility for adaptations to better reflect different contexts and understand sexual and reproductive health issues for many around the world.

## Data Availability

All data referred to in the manuscript is included in supplementary materials.

## Footnotes

**Twitter:** @ekpoks @YourLocalIDdoc @Dan_Wu_SESH @laleesay @JosephTucker @liannegonsalves

## Contributors

All authors wrote and edit the manuscript and approved the final draft. All authors confirm that they have contributed to this article and met the following three requirements: (a) they made significant contributions to the conception, design and implementation; (b) they drafted or revised the article for intellectual content; and (c) gave final approval of the submitted article.

## Funding

Financial support for this study was provided by the UNDP/UNFPA/UNICEF/WHO/World Bank Special Programme of Research, Development and Research Training in Human Reproduction, and the Academy of Medical Sciences and the Newton Fund (Grant number NIF\R1\181020). The funders played no role in the study design, data collection, interpretation of data, the writing of the report, or in the decision to submit the article for publication.

## Competing interests

The authors declare that they have no conflict of interests.

## Patient consent for publication

Not required.

## Acknowledgements

The authors would like to thank the following steering committee members for providing guidance throughout the different stages: Linda-Gail Bekker, Laura Lindberg, Annette Sohn, Emma Slaymaker and Pedro Nobre. We would also like to thank Martina Morris, Christopher Sengoga, Aleksandar Štulhofer, and Rocio Murad for their valuable contributions to the open call and hackathon. We acknowledge Lisa Atieno Omondi for the hackathon logistics arrangements. Thanks to the London School of Hygiene and Tropical Medicine (LSHTM) for general co-ordination, the UNDP/UNFPA/UNICEF/WHO/World Bank Special Programme of Research, Development and Research Training in Human Reproduction (HRP) for financial support and the African Population and Health Research Centre (APHRC) in Nairobi for hosting the hackathon.

## SUPPLEMENTARY MATERIALS

Supplementary file 1: Open call

Supplementary file 2: Judging criteria

Supplementary file 3: Hackathon guide

Supplementary file 4: Survey questionnaire for Delphi

Supplementary file 5: Draft survey instrument

Supplementary file 6: Implementation considerations

## Notes

### Competing Interest Statement

The authors have declared no competing interest.

## References

1. Edwards WM, Coleman E. Defining Sexual Health: A Descriptive Overview. Archives of Sexual Behavior 2004; 33(3): 189–95.

2. WHO. Defining sexual health: Report of a technical consultation on sexual health. 006 (accessed 13 Oct 2019).

3. Bajos N, Bozon M, Beltzer N, et al. Changes in sexual behaviours: from secular trends to public health policies.Aids 2010; 24(8): 1185–91.

4. Wellings K, Collumbien M, Slaymaker E, et al. Sexual behaviour in context: a global perspective. The Lancet 2006; 368(9548): 1706–28.

5. Hubert M, Bajos N, Sandfort T. Sexual behaviour and HIV/AIDS in Europe: Comparisons of national surveys: Routledge; 2020.

6. Stoller RJ. Sex and gender: The development of masculinity and femininity: Routledge; 2020. 7.

7. Shaeer O, Shaeer K. The Global Online Sexuality Survey (GOSS): the United States of America in 2011. Chapter I: erectile dysfunction among English-speakers. J Sex Med 2012; 9(12): 3018–27.

8. Kontula O. Sex Life Challenges: The Finnish Case. In: Wright JD, ed. International Encyclopedia of the Social & Behavioral Sciences (Second Edition). Oxford: Elsevier; 2015: 665-71.

9. Richters J, Badcock PB, Simpson JM, et al. Design and methods of the Second Australian Study of Health and Relationships. Sex Health 2014; 11(5): 383–96.

10. Mitchell KR, Mercer CH, Ploubidis GB, et al. Sexual function in Britain: findings from the third National Survey of Sexual Attitudes and Lifestyles (Natsal-3). Lancet (London, England) 2013; 382(9907): 1817–29.

11. Mercer CH, Tanton C, Prah P, et al. Changes in sexual attitudes and lifestyles in Britain through the life course and over time: findings from the National Surveys of Sexual Attitudes and Lifestyles (Natsal). The Lancet 2013; 382(9907): 1781–94.

12. Bajos N, Spira A, Ducot B, Messiah A. Analysis of sexual behaviour in France (ACSF): A comparison between two modes of investigation: Telephone survey and face-to-face survey. Aids 1992.

13. Kontula O. Sex life challenges: The Finnish case. 2015.

14. USAID. The Demographic and Health Surveys (DHS) Program. 2020. https://dhsprogram.com/What-We-Do/Survey-Types/DHS.cfm (accessed 18 May 2020). 15.

15. UNICEF. Multiple Indicator Cluster Surveys. 2020. https://www.unicef.org/statistics/index_24302.html (accessed 27 May 2020).

16. Slaymaker E, Scott RH, Palmer MJ, et al. Trends in sexual activity and demand for and use of modern contraceptive methods in 74 countries: a retrospective analysis of nationally representative surveys. The Lancet Global Health 2020; 8(4): e567–e79.

17. Tepper MS. Sexuality and disability: The missing discourse of pleasureSexuality and disability 2000; 18(4): 283–90.

18. Amaro H, Raj A, Reed E. Women’s sexual health: The need for feminist analyses in public health in the decade of behavior. Psychology of Women Quarterly 2001; 25(4): 324–34.

19. Kreps GL, Peterkin AD, Willes K, et al. Health care disparities and the LGBT population: Lexington Books; 2014.

20. Campbell S. Sexual health needs and the LGBT community. Nursing Standard 2013; 27(32).

21. Fredriksen-Goldsen KI, Kim H-J, Barkan SE, Muraco A, Hoy-Ellis CP. Health disparities among lesbian, gay, and bisexual older adults: Results from a population-based study. American journal of public health 2013; 103(10): 1802–9.

22. Tucker JD, Day S, Tang W, Bayus B. Crowdsourcing in medical research: concepts and applications. PeerJ 2019; 6: e6762.

23. World Health Organization, UNICEF. Crowdsourcing in health and health research: a practical guide: World Health Organization, 2018.

24. Tucker JD, Tang W, Li H, et al. Crowdsourcing Designathon: A New Model for Multisectoral Collaboration. BMJ Innovations 2018; 4: 46–50.

25. MIT Hacking Medicine. Health Hackathon Handbook. 2016.

26. Hasson F, Keeney S, McKenna H. Research guidelines for the Delphi survey technique. Journal of advanced nursing 2000; 32(4): 1008–15.

27. Humphreys TP, Brousseau MM. The sexual consent scale-revised: development, reliability, and preliminary validity. Journal of sex research 2010; 47(5): 420–8.

28. Pulerwitz J, Barker G. Measuring attitudes toward gender norms among young men in Brazil: development and psychometric evaluation of the GEM scale. Men and Masculinities 2008; 10(3): 322–38.

29. Fincham FD, Cui M, Braithwaite S, Pasley K. Attitudes toward intimate partner violence in dating relationships. Psychological assessment 2008; 20(3): 260.

30. Shah A. Hackathons Target Coronavirus. Participants tackle global problems from the shortage of ventilators to how to enforce social distancing. 2020. https://www.wsj.com/articles/hackathons-target-coronavirus-11586424603 (accessed 9 April 2020).

