## Supplemental Files for "Using a crowdsourcing open call, hackathon and a modified Delphi method to develop a consensus statement and sexual health survey instrument"

**Supplementary materials**

Supplementary file 1: Open call
Supplementary file 2: Judging criteria
Supplementary file 3: Hackathon guide
Supplementary file 4: Survey questionnaire for Delphi
Supplementary file 5: Draft survey instrument
Supplementary file 6: Survey Implementation considerations

**Supplementary file 1: Open call**

### Seeking feedback to develop a population-representative sexual health survey instrument:

#### An open call from the WHO

*Are you a sexual/reproductive health advocate or researcher? Passionate about sexual health in practice or research? The WHO and partners need your feedback on a survey instrument assessing sexual practices, behaviours, and outcomes.*

##### Background

To date, there is no standard, globally-recognized instrument to measure sexual practices, behaviours and sexual health-related outcomes. Instead, many population-representative surveys use their own items and domains, making comparisons and collaboration difficult. To encourage the inclusion of transparent and comparable sexual health-related measures on population-representative surveys, and in response to calls from leading sexual health researchers, **the WHO seeks to develop a standard instrument for assessing sexual practices, behaviours, and sexual health-related outcomes.** This instrument could then serve as a 'module' for use in national and sub-national data collection, as well as research.

The purpose of this open call is to solicit specific measures from a diverse range of advocates and researchers in order to create a standard sexual health research instrument.

##### Who can participate?

This call is open to anyone with professional interest, experience and/or expertise in sexual practices/behaviours and sexual health-related outcomes. This experience can be related to certain populations or the general population.

##### Why should I submit?

Your submission will help to develop this standard instrument for assessing sexual health practices, behaviours, and outcomes, and also encourage transparent and comparable sexual health items on population-representative surveys across the globe.

All submissions will be issued a commendation certificate to recognise participation. Exceptional submissions will be supported to attend a sexual health-related hackathon. The purpose of this consensus-building meeting will be to finalize the survey instrument, build capacity for global sexual health research, and plan next steps

human reproduction programme hrp research for impact
UNDP·UNFPA·UNICEF·WHO·WORLD BANK

World Health Organization

SOCIAL INNOVATION IN HEALTH INITIATIVE

LONDON SCHOOL of HYGIENE &TROPICAL MEDICINE

SESH Crowdsourcing For Health

#### Format and guidelines for submission

All submissions should be related to sexual and reproductive health. We are particularly interested in items that can be used in a broad range of settings and for the general population.

All measures and any comments can be provided in any of the six official WHO languages (these are Arabic, Chinese, English, French, Russian and Spanish): where possible, English is preferred.

Submissions can be:

1. Examples of existing survey instruments. Please provide as much information as possible on any instrument provided.
2. Examples of items or questions which could be used in survey instruments. Topics to consider include: life events including first sex, sexual partners, sexual practices, last sexual partner(s), sexual satisfaction, non-consensual sex, reproductive history/preferences, sexual health-related knowledge, and sex/sexuality-related social norms.

Important questions to consider include the following:

1. What is the optimal duration of recall for assessing these measures (last 3 months, 6 months, 12 months)?
2. What are the appropriate measures related to sexual health and sexual health practices?

Files can be uploaded as word documents or PDFs. All entries should be submitted via the website submission portal by **11:59 GMT on October 24th, 2019**

#### Follow up

All submissions will be issued a commendation certificate to recognise participation. Exceptional submissions will be supported to attend a sexual health-related hackathon. The purpose of this consensus-building meeting will be to finalize the survey instrument, build capacity for global sexual health research, and plan next steps.

The submissions will be reviewed by at least three independent individuals. Criteria for judging will be relevance to sexual health surveys (i.e., focused on sexual health population surveys), feasibility (i.e., is this practical and useful), and generalizability (able to be applied in a wide range of settings, across a general population).

#### Timelines

- **July-August 2019:** Establish a steering group to oversee the global call
- **September 2nd, 2019:** Launch the online call for submissions
- **October 24th 2019:** Deadline for submissions
- **Mid November:** Notification of submissions under consideration for hackathon participation.
- **November-December 2019:** Steering Committee review input and comments, determine relevance

- **January 2020**: In-person hackathon, hosted by an HRP Alliance hub (see below partners) providing experts from around the world 3 days to 'hack' together the final draft of the instrument
- **February 2020:** WHO review and finalize instrument

##### Submitting entries

All entries should be submitted via the website submission portal by 11:59 GMT on October 24th, 2019

##### Steering Committee Members

This global call is coordinated by a steering committee consisting of a global and multidisciplinary group of experts in sexual health:

Lianne Gonsalves (Co-Chair)- World Health Organization(WHO); Joseph Tucker (Co-Chair) - Social Entrepreneurship to Spur Health (SESH)Global; Lale Say – WHO; Megan Srinivas – University of North Carolina (UNC); Nathalie Bajos – French National Institute of Health and Medical Research (INSERM); Emma Slaymaker – London School of Hygiene and Tropical Medicine (LSHTM); Annette Sohn – The Foundation for AIDS Research (amfAR); Laura Lindberg - Guttmacher Institute; Pedro Nobre - World Association for Sexual Health; Linda-Gail Bekker – University of Cape Town/International Aids Society; Cesar Carcamo – Universidad Peruana Cayetano Heredia; Eneyi Kpokiri -SESH Global; Kaye Wellings – LSHTM; Boniface Ushie – African Population Health Research Center

##### Partner Organisations

##### London School of Hygiene and Tropical Medicine (LSHTM)

The LSHTM team has implemented 42 crowdsourcing events, including six global ones. Five randomized controlled trials from their team suggest that crowdsourcing can effectively engage communities and solicit effective entries. The LSHTM team was commissioned by the WHO HIV Department and the WHO Global Hepatitis Programme to write systematic reviews focused on diagnostics. LSHTM helped to launch the Social Innovation in Health Initiative (SIHI) in partnership with the WHO-hosted Special Programme for Research and Training in Tropical Diseases (TDR). In addition, they contributed to the 2018 guide to crowdsourcing in health and health research.

##### SESH

SESH, Social Entrepreneurship to Spur Health, is a partnership between universities focused on using crowdsourcing methods to improve health. SESH was founded in 2012 and has organized over 50 crowdsourcing challenge contests. SESH partnered with TDR to organize the Women Leaders in Global Health Challenge in 2018.

##### HRP Alliance Hub

The HRP Alliance for Research Capacity Strengthening is an initiative that brings together institutions conducting research in sexual and reproductive health and rights in collaboration with WHO regional and country offices. The HRP Alliance fulfils a mandate of supporting research capacity strengthening in low- and lower-middle income countries. An HRP Alliance Hub will host and co-lead the hackathon.

#### Supplementary file 2: Judging criteria

##### Judging entries for WHO/HRP sexual health challenge

Thank you for agreeing to be a judge for the Sexual Health Survey Instrument call for entries. We appreciate your time and support. Currently, there is no globally standardized instrument to measure sexual practices, behaviors, social norms and sexual health-related outcomes.

The purpose of this call is to create a sexual and reproductive health quantitative survey instrument that could be easily used in a broad number of settings, especially low- and middle-income countries. More information about the call is available here: https://www.who.int/news-room/detail/03-09-2019-seeking-feedback-to-develop-a-population-representative-sexual-health-survey-instrument

Please only consider the content of the entry, and disregard grammar, typographic or presentation errors/flaws. Please recuse yourself from judging entries with which you have a conflict of interest (for example, if you know the group who submitted a case, funded the project, or have been otherwise involved in the development or write up of the submission). Entries from which you have recused yourself should be noted with an R in the appropriate column on the scoring sheet.

Scores for each entry will be averaged and entries will be ranked. When scoring the entries, please use the following criteria:

1. **Relevance of the entry to inform a population representative survey instrument.** Does this entry provide insightful and innovative additions or improvements to the survey instrument such as new domains, implementation considerations, survey instruments, or creative ideas? Could this entry help the instrument become more inclusive of LMIC settings, vulnerable populations, or other groups?
2. **Participant's contribution in previous surveys and publications, experience in their field, and ability to contribute at the hackathon.** Does the participant's experience in their field and area of research prove that they would offer helpful and positive opinions, comments, and revisions at the hackathon?

[Rubric on next page]

| Criteria/Scores | 1-3 | 4-6 | 7-10 |
|---|---|---|---|
| Provided relevant and useful content (e.g. new domains or creative ideas, implementation considerations, survey instruments) | This entry provides little new content or considerations, and is not strongly related to creating a comprehensive, standardized instrument | This entry can provide somewhat relevant additions or considerations related to creating a comprehensive, standardized instrument | This entry contains robust content and highlights areas that could be improved or added to the instrument with a clear focus on several of the broad categories of the objectives |
| Participant's experience in the SRH field (including publications), cross national comparative experience, and contributions to development of the survey instrument, domains or new ideas | Participant has little to no experience in the field of suggested revision or addition; would not be a helpful contributor at the Hackathon | Participant has experience in the field of study highlighted in the submission; offers some insight in their submission as to their depth of knowledge and experience | Participant is clearly experienced in their designated field and shows through their submission that they would be able to provide an insightful and unique opinion at the Hackathon |

**Supplementary file 3: Hackathon guide**

### Hackathon Guide

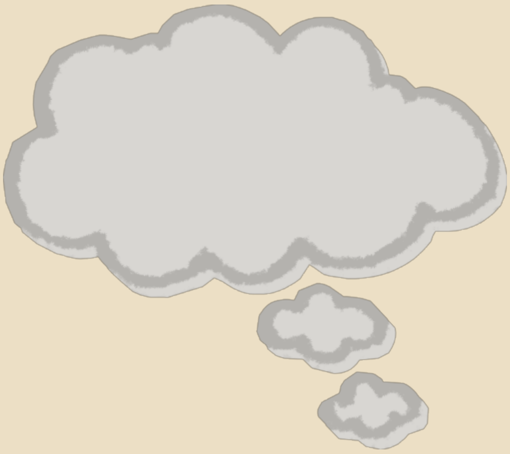

**14-16 January 2020**
**Nairobi, Kenya**

human reproduction programme hrp research for impact UNDP·UNFPA·UNICEF·WHO·WORLD BANK

World Health Organization

APHRC African Population and Health Research Center

LONDON SCHOOL of HYGIENE &TROPICAL MEDICINE

SESH Crowdsourcing For Health

Frequently Asked Questions… ...13-14

Meet the Facilitators ...................15-18

Meet the Participants .................. 19-28

Meet the Organizers…................... 29-33

Steering Committee ........................34

#### About the HRPHackathon

The UNDP/UNFPA/UNICEF/WHO/World Bank Special Program of Research, Development and Research Training in Human Reproduction (HRP) in partnership with the African Population and Health Research Center (APHRC), the London School of Hygiene and Tropical Medicine (LSHTM), and Social Entrepreneurship to Spur Health (SESH) will host a hackathon in Nairobi, Kenya. To our knowledge, this is one of the first health hackathons in Kenya. The purpose of the hackathon is to bring together sexual health experts, researchers, and those passionate about sexual health to develop a standardized instrument to assess sexual health, sexual behaviors, and sexual health-related outcomes. The goal is to develop a survey instrument that would be relevant in diverse global settings, especially low and middle- income countries. The hackathon will bring together participants to create a comprehensive survey instrument and suggest implementation techniques.

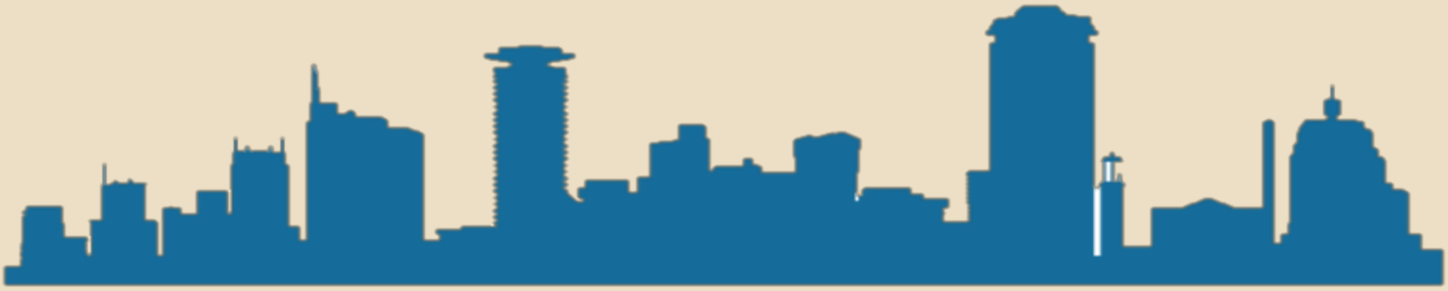

Nairobi Skyline

Adapted from www.trzcacak.com

### Why a hackathon?

A hackathon is a sprint-like event that brings together individuals with diverse backgrounds to solve a problem.[1] A hackathon can tap into participants' experiences and expertise to generate high quality outputs in a transparent and systematic way.[2]

A recent study conducted by LSHTM used a hackathon to identify exceptional ideas from the community to promote HIV testing among men who have sex with men in China.[3] Hackathons are one type of crowdsourcing in which a group of people solve a problem and then share solutions with the public.[4] Our sexual health survey hackathon will develop a sexual health survey instrument to measure sexual health practices, behaviours and related outcomes across different settings.

### Why isthis important?

The right to safe, consensual, and enjoyable sex is listed in the WHO's Constitution as a key component to achieving the "highest attainable standard of health." Sexual health is essential to one's overall health and wellbeing; however, to date, there is no all-inclusive, globally recognized assessment of sexual practices, behaviours and sexual health-related outcomes. Instead, some population-representative surveys use their own items and domains, making comparisons difficult. This underlines the need for a harmonized sexual health survey instrument to be used in global (especially LMIC) settings. At its most basic, an instrument could be a brief module embedded within another survey related to sexual health. A longer version of the instrument could also be a stand-alone survey instrument.

[1] *Health Hackathon Handbook - MIT Hacking Medicine. 2016. http://hackingmedicine.mit.edu/healthcare-hackathon-handbook/*
[2] *Tucker JD, Tang W, Li H, et al. Crowdsourcing designathon: a new model for multisectoral collaboration. BMJ Innovations 2018;* ***4****:46-50.*
[3] *Tang W., Wei C., Cao B., Wu D., Li K.T., Lu H., Ma W., (...), Tucker J.D. 2018. Crowdsourcing to expand HIV testing among men who have sex with men in China: A closed cohort stepped wedge cluster randomized controlled trial. PLoS Medicine, 15 (8), art. no. e1002645*
[4] *Tucker JD, Day S, Tang W, Bayus B. 2019. Crowdsourcing in medical research: concepts and applications. PeerJ 7:e6762 https://doi.org/10.7717/peerj.6762*

#### Problem Statement

Sexual health survey data are often difficult to compare across studies and between countries because survey instruments can be markedly different. In addition, many survey instruments were developed for use in high-income countries, without attention to undertaking similar research in low and middle-income countries (LMICs).

**What are the core elements of a sexual health survey instrument that could be used in diverse global settings?**

**How can we encourage uptake of the sexual health survey instrument in diverse global settings?**

The goal of this hackathon is to address these questions and create a sexual health survey instrument that will enable global sexual health research. Here we use the term "sexual health" to include reproductive health, recognizing that sexual and reproductive health are related. Many surveys of sexual health narrowly focus on life events and contraception, without appreciating sexual desire, sexual satisfaction, nonconsensual sex, and sexual rights.

Hackathon deliverables include a consensus statement on sexual health research, a tiered list of survey items and domains, and open access resources for sexual health research.

### Uses of the Survey

The purpose of this project is to create a sexual health survey to be used in a broad range of global settings. The following were collated from the pre-hackathon survey data:

**Survey types.** The main uses of the survey will be for population-representative research studies, trials related to sexual health, epidemiological research, implementation science, and others (humanitarian settings, primary care, and subgroups of interest).

**Participants.** The survey should be applicable to the following ranges of participants: participants who live in more conservative areas (oppose abortion/reproductive choice and have problems with sexual minorities) and people who live in more liberal areas (support reproductive choice and sexual minorities); participants without any sexual activity (especially younger and older groups) and people with higher number of sexual partners and greater sexual risk (e.g., some sex workers); participants from low-income countries and participants from upper middle-income countries; participants who have limited English language proficiency (B1 intermediate level).

**Administration.** We will focus on a survey administered with forms on mobile phones, tablets, or computers that **do not** have regular internet. Data can be collected offline and later uploaded. It will be interviewer administered with self-administration for selected components.

### How this works

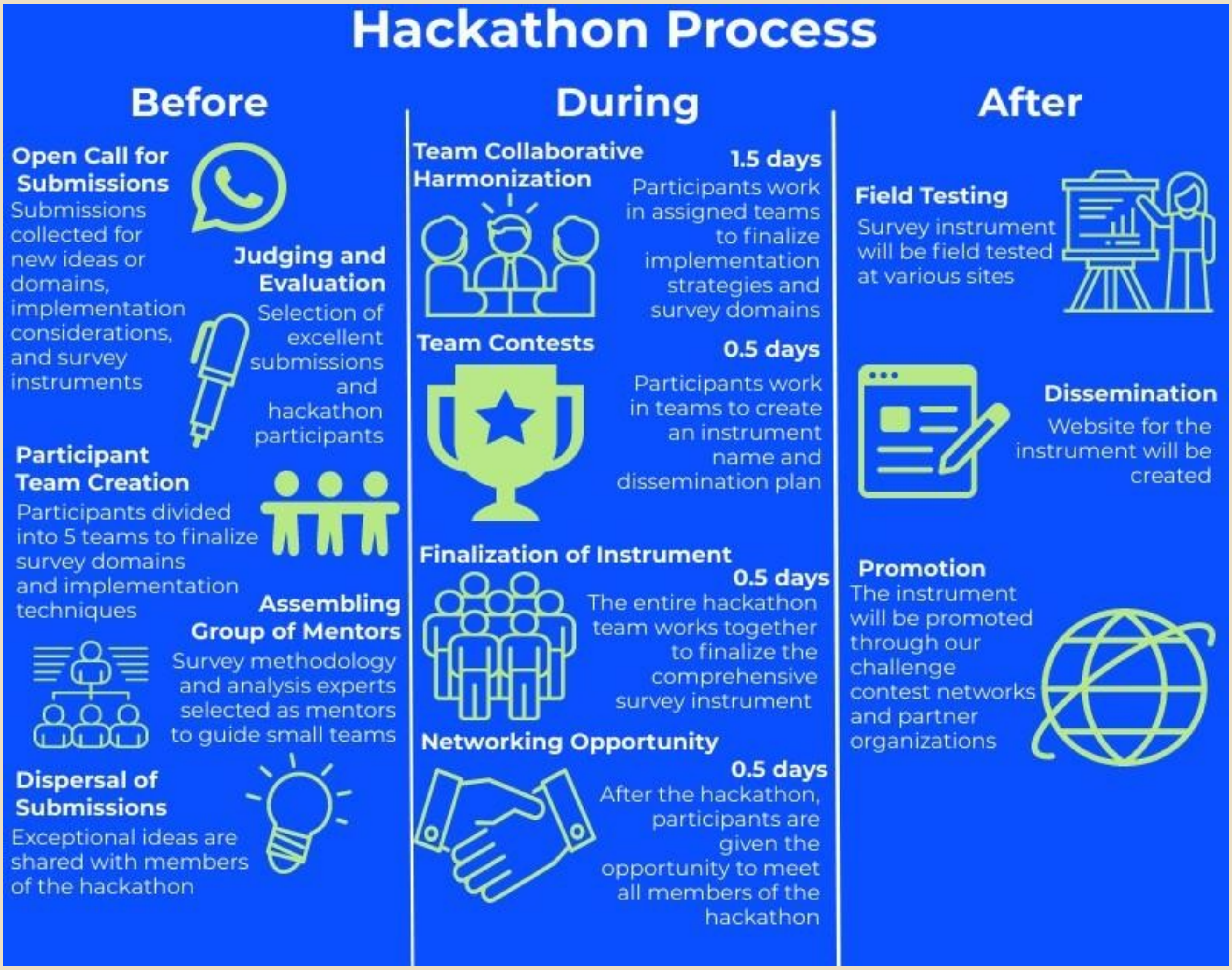

<u>Team Collaborative Harmonization and Team Contest</u>

**Team Collaborative Harmonization** – participants will work in teams for the first 1.5 days. Exceptional ideas identified through the challenge will be used to develop and finalize their designated survey domain or implementation technique on word documents. On the final half day, the teams will work together to synthesize all domains and implementation techniques creating one final harmonized survey instrument.

**Team Contest** – During the afternoon of day 2, each team will compete against one another to create the best survey name and dissemination methods. The best name and dissemination plan for the instrument will be determined by an adapted Delphi method.

Groups

Each group will have 1-2 organizers, one facilitator, and 3-4 participants. We have two floating facilitators and two floating organizers.

**Implementation Group (Facilitators – Noor Ani Ahmad/ Osmo Kontula; Organizer - Joe Tucker; Participants – Alice Welbourn, Chantal Smith, Christopher Sengoga and Kathryn O'Connell)** – interviewer training, stakeholder engagement, survey administration, protecting participants, data management, ethical review, translation, local support services, software, survey design issues

**General Information Group (Facilitator - Chima Izugbara; Organizer - Juliana Anderson; Participants – Michele Andrasik, Adesola Olumide, Wendy Norman and Nicole Prause)–** General health and disability, social demographics, & reproductive history/intentions, sexual partners

**Sexual Activity Group (Facilitator – Chelsea Morroni; Organizer - Megan Srinivas; Participants – Jennifer Toller Erausquin, Aleksandar Štulhofer and Martina Morris) –** Last sexual partners, sexual satisfaction, & sexual (dys)function

**Significant Events Group (Facilitator – Richard de Visser; Organizer - Eneyi Kpokiri; Participants – Soazig Clifton, Ariane van der Straten and Amanda Gabster) -** Significant life events related to sex/sexuality, first sex**,** & non-consensual sex

**Sexual Norms and Understanding Group (Facilitator - Georgina Yaa Oduro; Organizer Dan Wu; Participants – Rocío Murad, Amanda Gesselman and Martha Nicholson)-** Sexual practices (frequency and preferences), social norms around sex/sexuality, sexual health related knowledge, sexual rights

### Schedule

**Day 1**

| | |
|---|---|
| 8:00 | Facilitator/organizer pre-meeting (only for facilitators and organizers) |
| 9:00 | Welcome and introduction to the hackathon (user-centered design) |
| 9:30 | Review hackathon purpose, outputs, and structure; second Delphi survey |
| 10:00 | Collaborative survey harmonization |
| **12:00** | Lunch break |
| 13:00 | Collaborative survey harmonization |
| **15:00** | Coffee break |
| 15:30 | Collaborative survey harmonization |
| 18:00 | Report back from each group (3 minutes each) |

**Day 2**

| | |
|---|---|
| **8:30** | Recap from Day 1 |
| 9:00 | Collaborative survey harmonization |
| 11:30 | Submit progress on group work related to survey items, domains, andresources |
| **12:00** | Lunch break |
| 13:00 | Small team contest – each team will create a name (acronym)sand dissemination plan for the project |
| **15:00** | Coffee break |

**15:30** Reconvene with small teams to finalize name and dissemination plan

**17:00** Submit name and dissemination plan

**17:30** Foreign language break-out session (among French and Spanish speakers)

**18:00** Group dinner organized by APHRC (at Zen Gardens for all and group transport will be arranged)

#### Day 3

**8:30** Final Delphi survey

**9:00** Work collaboratively as a large group to finalize items

**10:30** Coffee break

**11:00** Plans for dissemination

**12:00** Lunch break

**13:00** Networking with facilitators and participants

**15:00** End of hackathon

#### Expectations androles

We are fortunate to have a tremendous group of individuals from all over the world join this hackathon. We received 139 applications from around the world to join. Each application was reviewed by three to five independent individuals and then final decisions were reviewed by the steering committee. Facilitators were recommended by one or more steering committee members, vetted by the organizing committee, and reviewed by WHO/HRP. As a result - **all facilitators, participants, and organizers joining us are the best of the best.** You all deserve to be here and we are delighted to have you!

**Facilitator** – These individuals will help to moderate discussion, resolve conflicts, and make sure that no one feels like an imposter. They are responsible for knowing their respective team members and getting those who are more quiet (or otherwise less likely to jump into discussions) to have their great ideas on the table.

**Organizer** – These individuals will help make sure that the group understands the goals/structure and stays on task. They have each reviewed peer-reviewed literature and responses to the open call related to the topics included in the group. This person will be a note-taker/scribe during the discussions.

**Participant** – These individuals bring their unique expertise and creative ideas to the hackathon. They could bring insight about survey design, development, or implementation.

#### Hackathon rules

Some things are not up for discussion. Below is a set of concepts that the hackathon steering committee agreed upon:

1) **Survey participant age.** The survey will focus on people aged 15 years and above, including ages 15-18 years old when feasible. We recognize the many adolescents would require a different survey, but we will not have sufficient time at the hackathonto develop a youth-focused version.
2) **English language.** We will focus on creating an English language survey instrument. Depending on the availability of proficient speakers of other languages (Spanish, French, Portuguese, Arabic), we will review selected non-English survey instrumentsas well.
3) **Overall survey duration of time needed to complete.** We anticipate that the overall survey time needed for completion would be as follows: 10 minutes (as an embedded module, highest priority items); 20 minutes (moderate priority items); 30 minutes (some priority). As a result, there will be three tiers of priority for domains – high, moderate, and some priority.
4) **Deliverables.** Hackathon deliverables will include the following: high-level consensus statement on principles (using an adapted Delphi method), a list of domains/items, open access resources for conducting sexual health research in global settings.
5) **Field readiness.** We will only consider survey items that have been used before, preferably in LMIC settings. We can include items that have only been used in high- income countries, but this would require more field testing. Items that are in development could be considered in a subsequent version of thesurvey.

#### Frequently AskedQuestions

How did you end up selecting the initial survey domains that were included in the HRP open call process?
The initial survey domains were generated at a Wellcome-Trust funded, WHO-convened consultation with experts in sexual health research in June 2019. More details about the process and outputs of this consultation are available in the Google drive folder “Hackathon - Collated Data by Group” in the subfolder called “WHO London materials.”

How were responses to the open call evaluated?
All responses to the open call were evaluated using a multi-stage process including determination of eligibility, judging, and steering committee assessment. Each individual submission was reviewed by five independent individuals and final decisions were made by the steering committee. Then the organizing group went through all responses to distill them into survey items, domains, and suggestions worthy of further consideration.

Will the WHO or others be supporting implementers to use this survey?
While more resources are helpful, we do not anticipate that the engine here will be new resources for sexual health research in LMICs. We must develop a survey instrument that is useful and feasible in a resource-constrained context.

Are there any survey items that are off limits?
The steering committee has established several rules (see page 12) that establish constraints. There are also some items discussed at the WHO London meeting which are well covered elsewhere and will not be the focus here. For example, survey items on treatment seeking and treatment access were deemed to be of lesser importance. In addition, it will be important for groups to prioritize their discussions of subtopics in order to best use the limited time available.

Why has this not been done before and how is this hackathon different?
Some have attempted to create a harmonized sexual health survey instrument, but these have not been widely used. Previous attempts have been stymied by limited input from LMIC researchers and less sexual and reproductive health research in LMICs. We believe that the hackathon format will leverage the collective strengths of participants, especially those with rich LMIC research experience. We will pro-actively identify areas where consensus may be impossible and draw on the creative ideas that have already been generated by the open call.

What are the most common sexual problems worldwide?
We do not have data to infer about the most common sexual problems on a global basis. We know from population-based representative studies in high-income countries that imbalances in sexual desire between a person and their partner are common. Yet there have been few similar studies in LMIC settings. Further research on sexual desire and sexual satisfaction in LMIC settings are needed.

Do I need to bring my laptop?
A laptop is not required, but it would be helpful to bring a personal computer.

Will we have wifi?
The conference venue will have wifi, but we encourage everyone to focus on the hackathon as much as possible.

What are online resources available?
The below items have been assembled by our organizer group:

HRP Hackathon draft documents (distilled from open call):
https://docs.google.com/document/d/1w2f1AZ1hF9D5BYJP8eQjvVKZlDtZgw2W1gW0YsnEz gY/edit?ts=5df10229

HRP Collated Data link: https://drive.google.com/drive/folders/1CThBDL_JtCBdONPXSiSszznoUkt4L5Uv

How do we prioritize ideas?
We will put survey domains into several categories: Tier 1 (high priority, for an overall 10- minute survey), Tier 2 (moderate priority, for an overall 20-minute survey), Tier 3 (some priority, for an overall 30-minute survey), parking lot (contentious, need for research), and other (contentious, no need for research).

#### Meet theFacilitators

**Osmo Kontula**

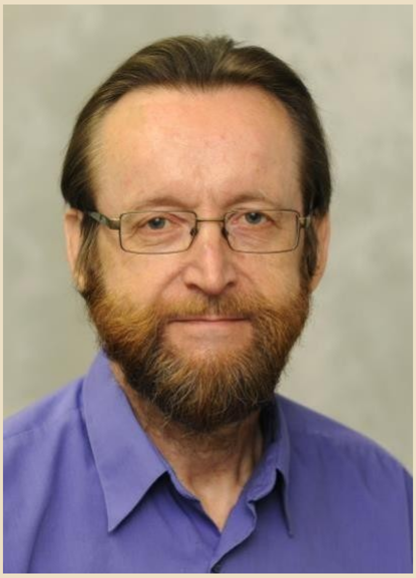

Osmo Kontula, Ph.D., is a Research Professor at the Population Research Institute of the Family Federation of Finland. He has authored around 400 publications, of which more than 50 are books. These include over 30 books in sexual health and sexology issues. Kontula has conducted, for example, FINSEX study that includes five nationally representative sexuality surveys since the 1990s in Finland. In addition, there has been a number of qualitative studies of sexuality issues. Fields of expertise include sexology, sexual science, sex research, sexual and reproductive health, cultural differences in sexual issues,sex education, adolescent sexuality, couple relationships, divorces, family, population and sexual policies, fertility, and demographic behavior.

Osmo Kontula is an Associate Editor of the Journal of Sex Research (JSR), a Member of Advisory Committee and a Chair of Sexuality Education Committee in the World Association for Sexual Health (WAS) since 2013, a full member of International Academy of Sex Research (IASR) since 1996, and a Past President of the Society for the Scientific Study of Sexuality (SSSS) and Nordic Association for Clinical Sexology (NACS). He has received Gold Medal from the European Federation of Sexology (EFS) in 2010.

Richard de Visser

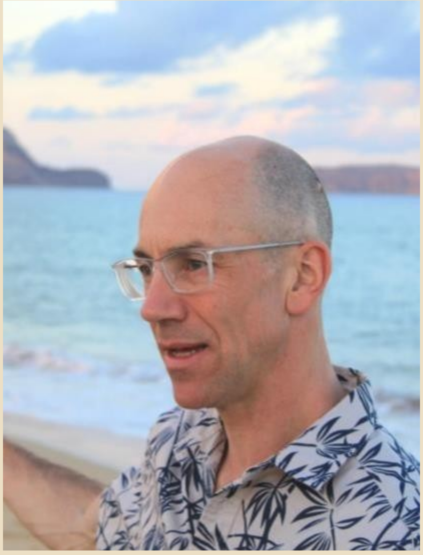

Richard de Visser teaches at Brighton & Sussex Medical School and in the School of Psychology at the University of Sussex. His research interests span a broad range of topics in health and social psychology, including: sexuality and relationships; use of health services; gender and health; alcohol use; and cross- cultural analyses. He has expertise in qualitative and quantitative methods, intervention studies, and mixed-methods designs. He has published over 120 articles in peer-reviewed journals, and is an author of the textbooks “Psychology for Medicine” and “Psychology for Medicine and Health Care”. Dr. de Visser’s key research activates in the domain of sexual health include the Australian Study of Health & Relationships, a population-representative study of 20,000+ people, which is now in its third iteration.

Cath Mercer

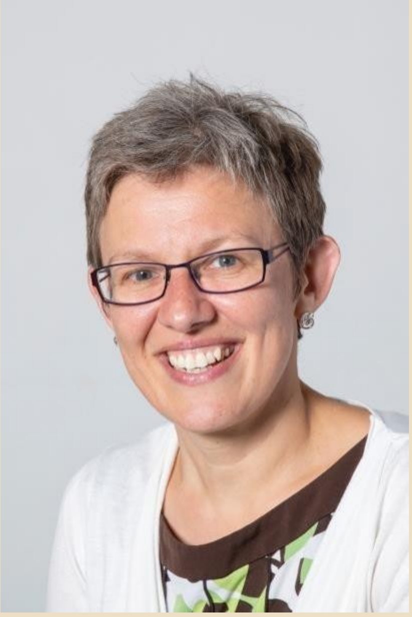

A statistician and demographer by training, Cath Mercer is Professor of Sexual Health Science in the Institute for Global Health at University College London. Cath is best known for leading Britain's National Survey of Sexual Attitudes and Lifestyles, Natsal, one of the world's largest and most reliable sources of scientific data on sexual behaviour, its drivers and health consequences. The Natsal data have been used to evaluate and inform a number of public health interventions and policy, e.g. HPV vaccination, teenage pregnancy, and sex and relationship education. Aside from leading impactful

research, Cath's expertise lies in developing and employing robust methods that advance the scientific study of sexual behaviour as well as sexual health more broadly, including measuring sexual behaviour in a range of hard-to-reach groups in diverse settings, as well as providing the general population perspective. She has a particular interest in developing methods that seek to go beyond the individual to better understand sexual risk as well as sexual well-being more broadly. Cath has published >200 papers in the field of sexual and reproductive health and has secured >£30M in competitive grant income. She is also an Associate Editor for the BMJ journal Sexually Transmitted Infections.

Nathalie Bajos

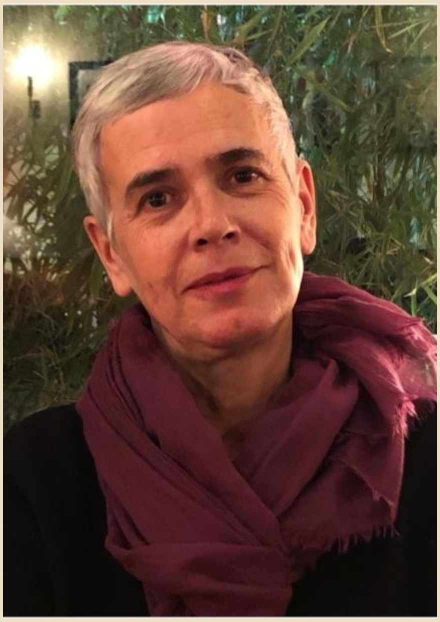

Nathalie Bajos is a sociologist and a demographer. She is the research director at the National Institute for Health and Medical Research (INSERM) in Paris, France. Specialist in gender and sexuality issues, she is responsible for major quantitative and qualitative national surveys on sexuality and sexual health in France (1992, 2006, 2020).

She has participated in numerous international comparisons on these topics. She was also responsible for the fight against discrimination and access to rights for the Human Rights Defender in France between 2015 and 2018. Her current work also focuses on social inequalities in health from gender and intersectional perspective.

Chelsea Morroni

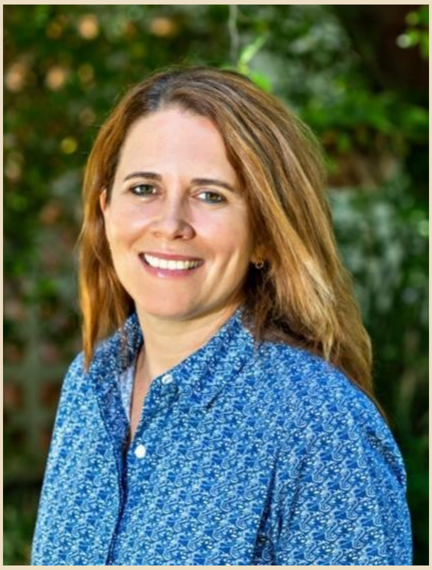

Chelsea Morroni is an epidemiologist and sexual and reproductive health (SRH) doctor. She is a Reader in International Sexual and Reproductive Health at the Liverpool School of Tropical Medicine, based full-time in Botswana; an honorary Professor in Public Health at the University of Cape Town (UCT), and holds research/clinical positions at Botswana- Harvard AIDS Institute and Botswana-UPenn Partnership.
Chelsea is Deputy Director of the UK Faculty of Sexual and Reproductive Healthcare's (FSRH) Clinical Effectiveness Unit in Edinburgh, and consults for the WHO, British Pregnancy Advisory Service, Margaret Pyke Trust and International AIDS Society.
She has 20 years of experience conducting clinical, health- service, and community-based research and doing policy/advocacy work on SRH in Southern Africa, focusing on contraception, abortion, STIs, and HIV-SRH integration. She is a volunteer advisor to the Botswana Ministry of Health, and is actively involved in clinical care and training and mentoring of healthcare providers in Botswana.

Noor Ani

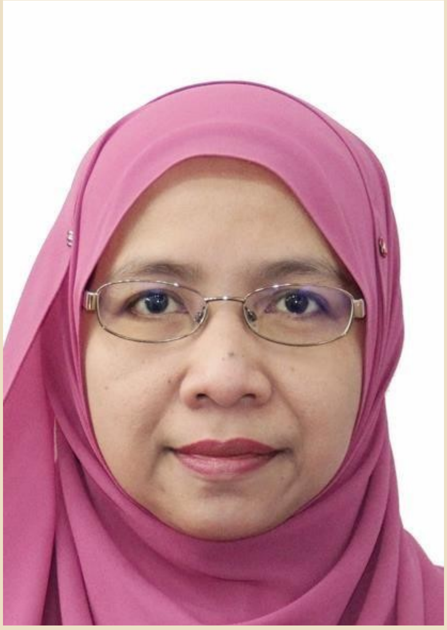

Noor Ani Ahmad is a Public Health Medicine Specialist and Head Centre for Family Health Research at the Institute for Public Health, Ministry of Health Malaysia. Dr Noor Ani has completed her medical degree and Master's in Public Health from University of Malaya, Malaysia. She has been involved in the implementation of national survey since 2005 and had led or been involved in the National Health and Morbidity Survey (NHMS), nation-wide population-based survey, since 2010. The surveys incorporated various scopes including sexual- reproductive health topics. She was also the Coordinator of the Malaysia Global School-based Student Health Survey in 2012, which included a topic on sexual activity of the adolescents.

She's currently the advisor for the planning of the NHMS.
Dr Noor Ani is currently the alternate Chairperson for the Ministry of Health Research Review Board and member of the R&D Review Board for the Ministry of Energy, Science, Technology, Environment and Climate Change, MESTEC, Malaysia. She has interest in the areas related to sexual reproductive health, mental health and disabilities. She has authored more than 50 articles including those related to sexual reproductive health, mental health and disabilities.

Georgina Yaa Oduro

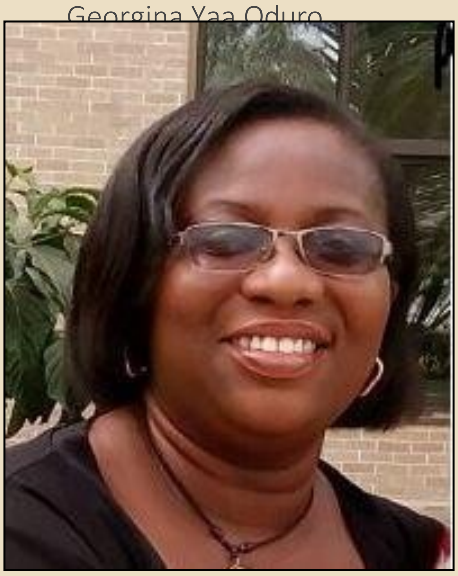

Dr. Georgina Yaa Oduro is a senior lecturer with the Department of Sociology and Anthropology, University of Cape Coast, Ghana. She holds a PhD degree in Sociology of Education from the University of Cambridge, UK. Master's degree from the University of Westminster- London, UK and a First Degree in Sociology and Political Science from the University of Ghana, Legon.

Dr. Oduro's PhD focused on Gender relations, sexuality and HIV/AIDS education from a youth culture perspective. This study has informed her research interest in Gender Issues, Violence, Sexuality, Youth Cultures, Popular Culture and Race and Ethnicity. She also has expertise in qualitative research methodologies. She has won a number of awards and fellowships with the latest being the Takemi Fellowship in International Health (2016-2017) at the Harvard T. H. Chan School of Public Health (Harvard University, Boston, USA) during which she researched child prostitution in Ghana. She has further conducted research on Abortion and the sexual lives of vulnerable populations including street youth. Dr. Oduro has a number of publications to her credit with some featuring in the Palgrave Handbook for Sexuality Education (2017) as well as the Routledge International Handbook for Sex Industry research (2019). Dr. Oduro is the current Director for the Centre for Gender Research, Advocacy and and Documentation (CEGRAD) of the University of Cape Coast in Ghana.

Chima Izugbara

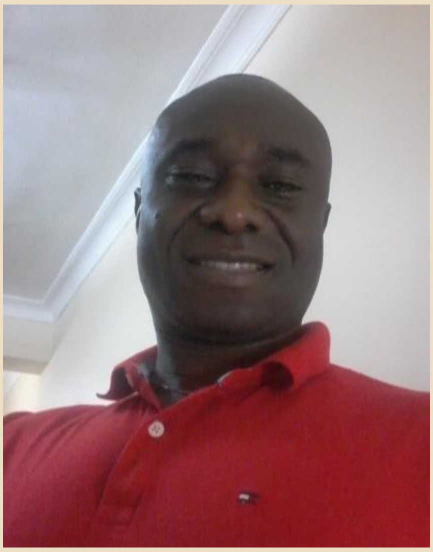

Chima Izugbara is currently Director, Global Health, Youth & Development at the International Center for Research on Women (ICRW), Washington DC. Prior to joining ICRW, he directed the Population Dynamics and Reproductive Health Program at the African Population and Health Research Center (APHRC), after leading the institution's Research Capacity Strengthening (RCS) Division for nearly a decade. A professor-at-large at the School of Public Health, University of the Witwatersrand, South Africa, Dr. Izugbara has taught in universities in multiple continents. A leading international scholar and researcher on gender, youth, sexuality and maternal, sexual, and reproductive health, Dr.

Izugbara holds two PhDs, the first in Health Anthropology and the second in Social Work (Gender, Health, and Development)
from the University of Gothenburg, Sweden.

#### Meet theParticipants

Aleksandar Štulhofer

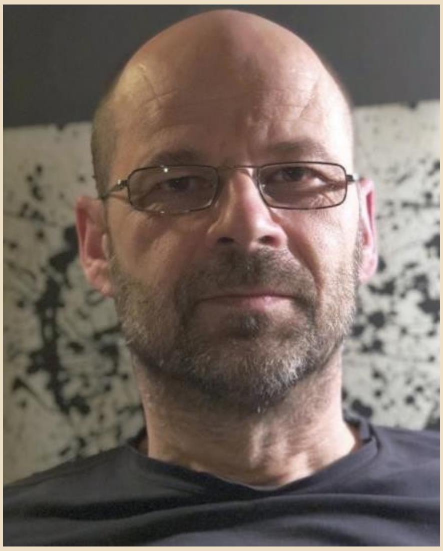

Aleksandar Štulhofer is professor of sociology and head of Sexology Unit at the Department of Sociology, Faculty of Humanities and Social Sciences, University of Zagreb, Croatia. He has published internationally on the epidemiology of sexual health, pornography and sexual socialization, hypersexuality, sexual satisfaction, emotional intimacy and sexual well-being, HIV risks and sexual risk taking, school-based sexuality education, and sexuality in older age. In the 2005-2016 period, Dr. Štulhofer served as short-term consultant for the WHO in the area of HIV surveillance. He was a full member of the International Academy of Sex Research (until 2019); currently he is a member of the Scientific Committee of the European Federation of Sexology, an Affiliated Faculty of the Kinsey Institute (since 2008), and a member of the European Society for Sexual Medicine. Dr. Štulhofer serves on the editorial board of the journals Archives of Sexual Behavior, Journal of Sex Research, and Sexuality and Culture. In 2016, he was awarded a Gold Medal from the European Federation of Sexology for contribution to European Sexual Health. His most recent research projects focus on longitudinal assessment of ties between adolescents' pornography use and well-being, healthy sexual aging in individuals and couples from five European countries, and links between sexual abuse and sexual health disturbances in adolescence.

Amanda Gabster

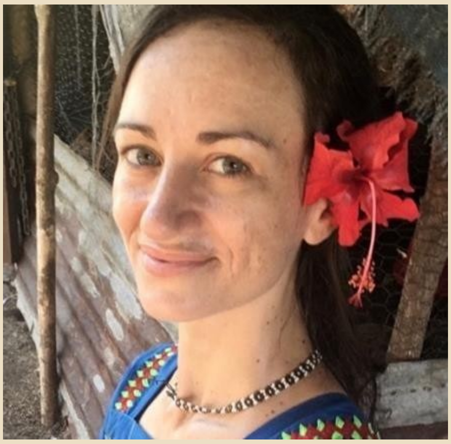

Amanda has been working in sexual health since 2009. The earlier years were devoted to sexual and reproductive health education with adolescents in rural communities of Panama through a non-profit she directs. In 2012, she began working at the Gorgas Memorial Institute in Panama City, in HIV and STI epidemiology, focusing mainly on adolescent populations since. In 2017, Amanda started her Ph.D. program within the Faculty of Infectious and Tropical Diseases of the London School of Hygiene and Tropical Medicine; her supervisors are Dr. Philippe Mayaud and Ben Cislaghi. Amanda´s current research focuses on STI epidemiology, especially social determinants of STI acquisition among adolescents of the Ngäbe-Buglé Indigenous region in Panama.

Martha Nicholson

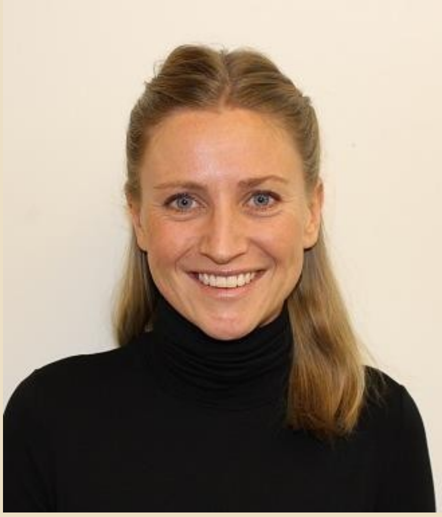

As the Evidence, Insights and Evaluation Advisor in the Strategic Evidence Team at Marie Stopes International (MSI), her role involves generating and disseminating evidence on how the organization can best respond to reproductive health needs of low and middle-income populations. During her work for Marie Stopes, she has designed a study on abortion-seeking behavior amongst rural populations in South Africa, supervised a literature review on the feasibility of using telemedicine for medical abortion care, and developed an inter-organization scale for safety of abortion service delivery. She is currently advising on an evaluation of Value Clarification and Attitude Transformation (VCAT) workshops on contraceptive and abortion client experiences in Ethiopia. She is also advising on an evaluation of behaviorally-informed job aids on the impact on Marie Stopes Uganda clients' capacity to achieve their reproductive aspirations through continuing on contraceptive methods or switching to a new method. Before starting work at MSI, she worked as an analyst in the department of Data Analytics and Epidemiology at Mapi Group consultants (part of ICON Sweden). She has a background of working, studying, volunteering and researching in the field of SRHR and affordability with Marie Stopes South Africa and RFSU Sweden

**Rocio Murad Rivera**

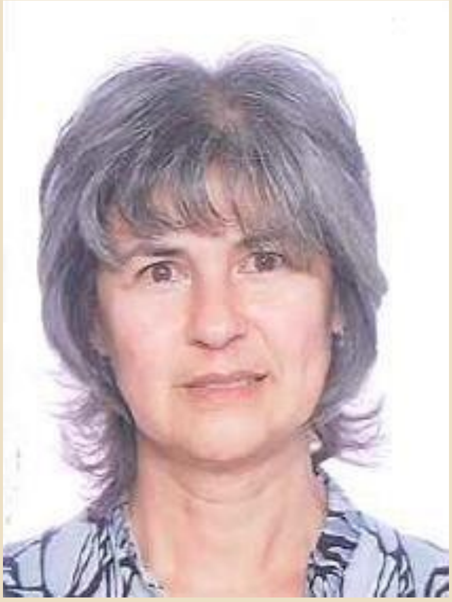

Rocío Murad Rivera, I am Colombian, statistician of the National University of Colombia with in-depth studies in Demography of the Latin American and Caribbean Demographic Center. My areas of interest are: sexual and reproductive law and sexual and reproductive health; teenage pregnancy; unmet need for contraceptive methods; demography; probabilistic population sampling; displacement and forced migration.

I have worked in Profamilia since 1987, where I am Coordinator of Sociodemographic Research. I have participated in the planning, development and analysis of multiple population surveys, among which I highlight the National Demographic and Health Surveys of Colombia (ENDS) from 1990 to 2015 and in the surveys on sexual and reproductive health of women in situations of displacement through the conflict of 2000, 2005 and 2011. I am currently participating in different research initiatives on the identification of health and sexual and reproductive health needs of the Venezuelan migrant population and I hope to start an investigation on HIV risk and the impact of migration soon sexual practices

Ariane van der Straten

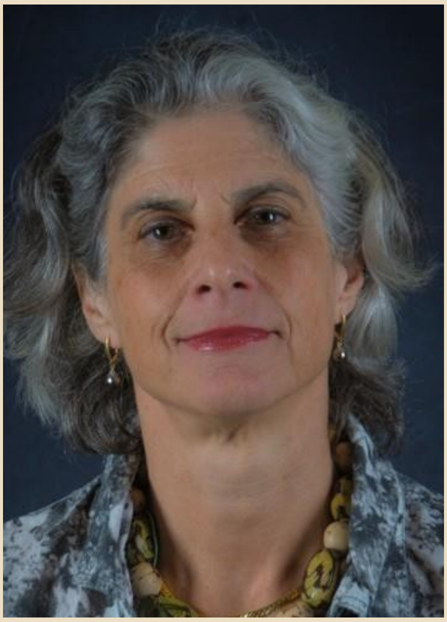

Ariane van der Straten received her PhD in molecular biology at the University of Brussels, (Belgium), and her MPH at the Johns Hopkins University, Baltimore. She is the Director of the San Francisco-based Women's Global Health Imperative (WGHI) program within RTI, and a Senior Fellow at RTI International. She is a Professor at the UCSF School of Medicine, Center for AIDS Prevention Studies and serves as the Chair of the Behavioral Research Working Group of the NIH-funded Microbicide Trial Network (MTN) and HIV Prevention Trial Network (HPTN).
Dr. van der Straten has over 25 years of experience conducting socio-behavioral and biomedical research for HIV prevention in women, including preclinical and phase I to phase III trials evaluating short and long-acting HIV prevention approaches (topical, oral, injectable, or implantable), and multi-purpose prevention technologies (MPT) for HIV and pregnancy prevention. Her current interests include the interplay of prevention technologies and behavior in the context of acceptability and adherence research. To that end she is leading studies that focus on understanding product preferences and attributes most suitable to end users and gatekeepers (e.g., health providers, male partners), using traditional qualitative and quantitative methods and marketing research approaches to better understand end-user choice and behaviors. With a team of engineers and laboratory scientists are RTI, she is also leading the development of end-user informed long-acting delivery approaches for HIV prevention and MPTs in the form of biodedgradable implants.

Wendy Norman

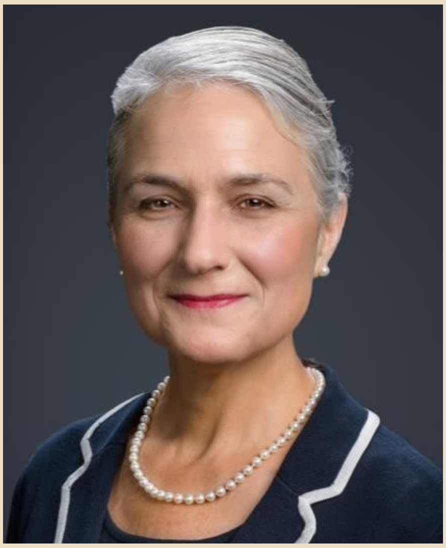

Dr. Wendy V. Norman, MD, MHSc, CCFP, FCFP, DTM&H, is a family physician-researcher with over 30 years clinical experience in sexual health and family planning clinics. She holds the Chair in Family Planning Public Health Research from the Canadian Institutes of Health Research and Public Health Agency of Canada; is an Associate Professor in the Faculty of Medicine, University of British Columbia, in Canada; and an Honorary Associate Professor in the Faculty of Public Health and Policy at the London School of Hygiene & Tropical Medicine in the UK. Dr. Norman fielded the BC Sexual Health Survey, and is working with the Government of Canada to implement the Canadian Sexual Health Survey designed by the team she has led. In 2015 Dr. Norman was awarded the prestigious Guttmacher Darroch Award for advancing reproductive health policy research. She founded and leads the national collaboration: The Canadian Contraception and Abortion Research Team. (www.cart-grac.ca)

Alice Welbourn

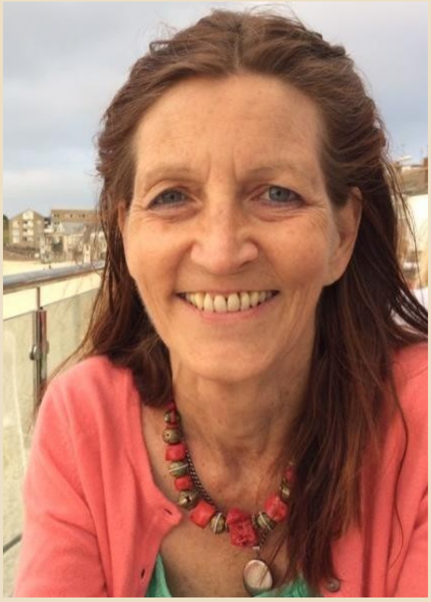

Alice Welbourn PhD FRCOG (Hon) is the Founding Director of Salamander Trust, a social anthropologist and a trainer, researcher, writer and advocate on, and of, SRHR and gender justice in the context of HIV. She lived in and worked for NGOs in rural Kenya and urban Somalia for most of the 1980s before many short-term training and learning experiences across rural East, West and Southern Africa. These enabled her to gain at first hand both depth and breadth in understanding gender, power, intersectionalities, and health, through participatory approaches to research and to community social norms change in diverse contexts. Diagnosed with HIV in 1992, she developed the Stepping Stones community training programme which produces multiple positive outcomes, including improved SRHR, and reduced VAWG (a significant barrier to SRHR) in the context of HIV. As chair of the International Community of Women living with HIV (ICW) from 2002-7, she advocated for the SRHR of women living with HIV at global level. In 2013 Salamander Trust was commissioned by WHO HRP to conduct a global values & preferences study of the SRHR of women living with HIV. This informed the WHO 2017 Guideline on this topic and, most recently the Checklist for its implementation, also published by WHO. The work on participatory research continues through the ALIV[H]E framework, for which Alice was co-PI, commissioned by UNAIDS. Alice is a regular adviser to UNAIDS, WHO, and other UN entities. See Salamander's latest newsletter for more information about Salamander's various other programmes, all of which relate closely to SRHR.

Nicole Prause

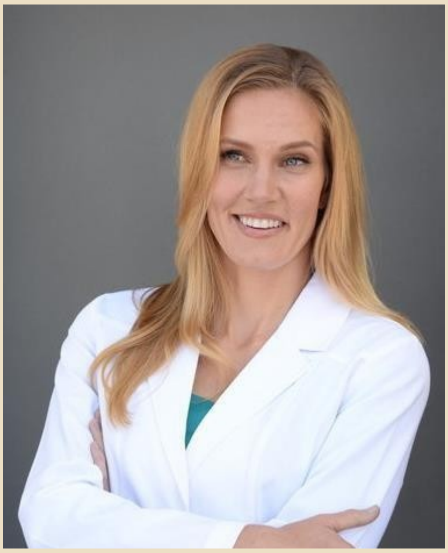

Nicole Prause, PhD is a sexual psychophysiologist who studies sexual decision making in experimental, laboratory research. Her research identifies predictors and methods for altering the sexual desires and motivations that drive sexual behaviors. She works with the University of Pittsburgh and the University of Nebraska- Lincoln on grant-funded, cutting-edge protocols that include sexual partners and genital stimulation to understand these factors.

Adesola Olumide

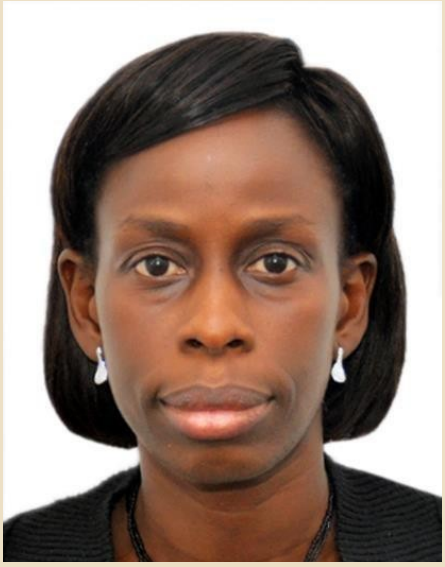

Adesola Olumide is a Senior Medical Research Fellow and Consultant Physician at the Institute of Child Health, University of Ibadan and University College Hospital, Ibadan, Nigeria. She coordinated the Adolescent Health component of the MPH Child and Adolescent Health (CAH) course run by the Institute of Child Health from 2008 until August, 2017. Her research interests include the epidemiology and risk factors for non-communicable conditions and health risk behaviors among adolescents. She has served as Principal Investigator (PI)/ Project Lead and Co-PI on a number of projects. She has experience working on studies focusing on the sexual health problems of diverse populations of adolescents and young people including in-and out-of-school, and very young adolescents. Some of these include a study of the HIV/AIDS knowledge and sexual practices of hearing- impaired students in Ibadan, Nigeria; the multi-country Well-being of Adolescents in Vulnerable Environments (WAVE) study, and an exploration of the predictors and economic costs of selected health-risk behaviours (including risky sexual practices) of adolescents in Ibadan, Nigeria. Adesola has a keen interest in the use of electronic media to reach adolescents and young people with health interventions. She is currently leading a project that aims to improve parents' capacity to communicate sexual and reproductive health information to their pre and early adolescents. Adesola was invited to serve as a Commissioner on the WHO- UNICEF-Lancet Commission on Child Health and Well-Being.

The Commission is charged with the responsibility of developing a report that highlights the importance of children and adolescents as key to the Sustainable Development Goals. She serves on the National Adolescent Health Technical Working Group in Nigeria and in this capacity has been involved in developing and revising key adolescent health documents and training manuals for the Federal Ministry of Health including the adolescent health policy which is currently undergoing revision. Adesola is the Secretary of the Society for Adolescent and Young People's Health in Nigeria (SAYPHIN).

Jennifer Erasquin

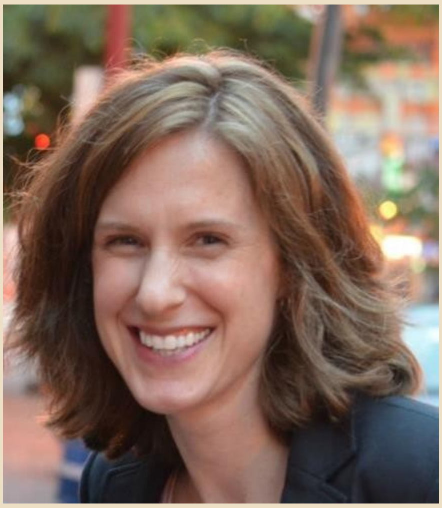

Dr. Erausquin is a social epidemiologist with the Department of Public Health Education at the University of North Carolina at Greensboro. She earned her undergraduate degree from the University of Michigan and her MPH and PhD from the UCLA Fielding School of Public Health. She was a predoctoral fellow of the California Center for Population Research and the UCLA AIDS Research Training Program, receiving training in demography and epidemiology. She went on to complete a postdoctoral research fellowship at the Duke Global Health Institute. Dr. Erausquin's research focuses on the intersections of gender, race/ethnicity, and socioeconomic factors as determinants of sexual and reproductive health. She is a quantitative methodologist with expertise in sampling hard-to- reach populations and analyzing large survey data sets, and a substantive expert in structural approaches to understanding and eliminating race- and gender-based disparities in health. She has made significant contributions to the field of sexual health, notably around the health of female sex workers. In 2015, her work on police practices and sex worker health was selected by UNAIDS' HIV This Month as top newsworthy research. In 2018, she served as co-editor of a compiled volume, Global Perspectives on Women's Sexual and Reproductive Health Across the Lifecourse (Springer, 2018).

Amanda Gesselman

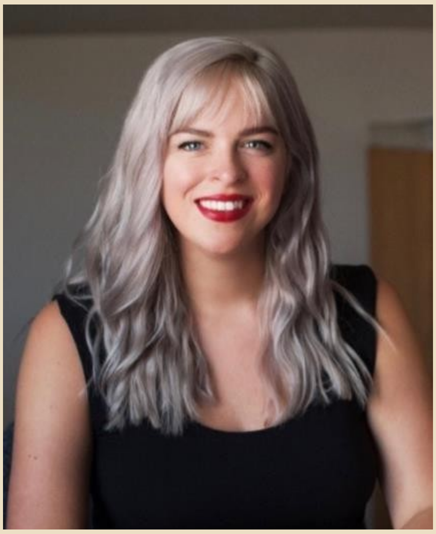

Dr. Amanda Gesselman is the Associate Director of Research, Anita Aldrich Endowed Research Scientist, and Head of the Research Analytics and Methodology Core at the Kinsey Institute at Indiana University, and is a Research Fellow at the Rural Center for AIDS/STD Prevention. She is a social–developmental psychologist, methodologist, and statistician, and has been working in the area of sexuality and health for nearly a decade.
Her research focuses on the interweavings of psychology, sexuality, and health in intimate relationships, with specific focus on how partners impact one another's mental and physical health, as well as how outside forces (e.g., technology, social stigma, societal norms) influence health and behavior.

Christopher Sengoga

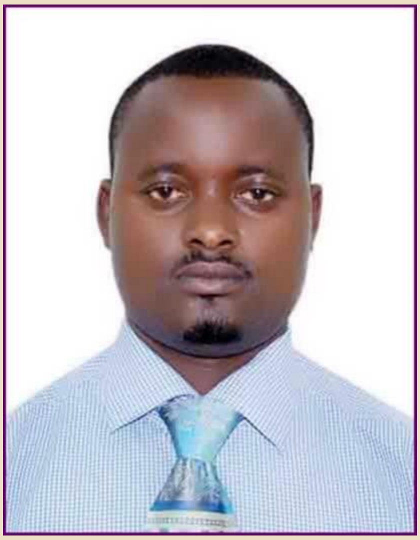

Mr. Christopher is currently pursuing a Master's degree in Sexual Reproductive Rights in Africa at the University of Pretoria in South Africa and holds Bachelor's degree in Laws (LLB) from the National University of Rwanda. Holds certificate on Sustainable Development and Human Rights Law at the University of Antwerp in Belgium; Crime, law and society from Sheffield University, UK. He hasworked with RR Associates & Co. Advocates as a Legal Associates; Great Lakes Initiative for Human Rights and Development (GLIHD) and worked with Oxfam's as Gender Justice Lead and currently work with Health Development Initiative as the Head of Human Rights and SRHR. The current areas of interest include; assisting vulnerable and poor women to access safe abortion, litigation on the right to health. In addition, he has raised literacy on SRHR among CSOs members; Healthcare providers; Journalists; Law enforcement authorities; Advocates from Rwanda Bar Associations; 10 University gender ministers and policy makers. He has been consulted for various tasks on SRHR by CSOs, government institutions (Rwanda Law Reform Commission, Ministry of Justice, Gender and Family Planning and Ministry of Health) and INGOs across East Africa. He has led an advocacy movement that contributed to the discrimination of abortion, same sex relationships and sex work in 2012 and 2018 penal review respectively in Rwanda. HDI has led a research on qualitative study on abortion of women convicted in prisons for the crime of abortion looking at the causes, the practices and the consequences. He has contributed to the release of women who were pardoned by the President of the Republic of Rwanda in 2019. Currently, Christopher is working on a research paper on the "Role of Catholic Church on abortion in Rwanda. He has broad knowledge on legal and policy analysis, GBV and broad range of experience in SRHR of women.

Soazig Clifton

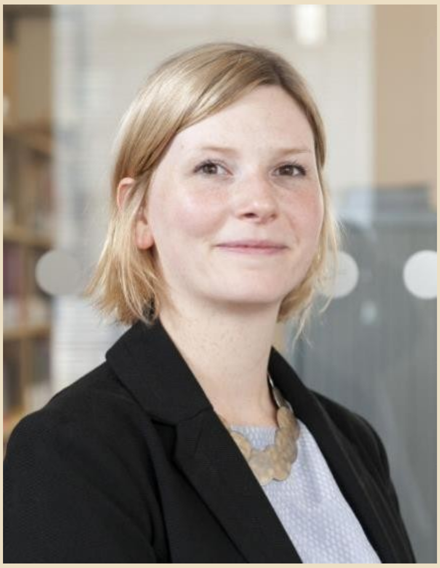

Soazig is a survey methodologist and epidemiologist based in London, UK. She has over 13 years' experience in the design, delivery, analysis, publication and dissemination of large-scale general population health research, with a focus on sexual behaviour and sexual health in Britain. She has been part of the core team on the British National Survey of Sexual Attitudes and Lifestyles (Natsal) since 2008, and leads on questionnaire development and testing, and survey implementation. She teaches questionnaire design, with a particular focus on sensitive topics, and is frequently invited to advise on questionnaire design for surveys nationally and internationally. She is jointly employed by University College London and NatCen Social Research.

Michele Andrasik

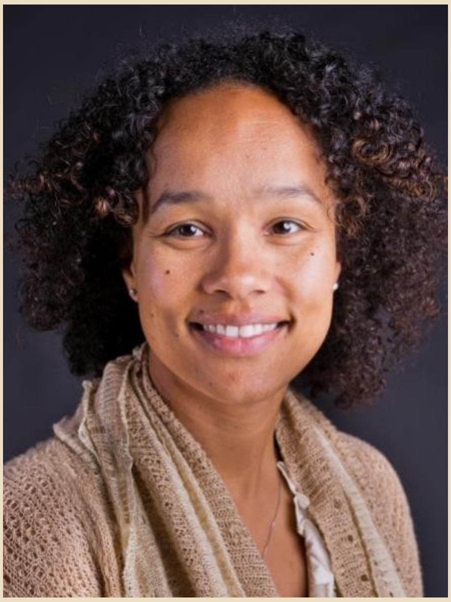

Michele Andrasik is a clinical health psychologist. She is the Director of Social and Behavioral Sciences and Community Engagement for the Fred Hutchinson-based HIV Vaccine Trials Network (HVTN), Senior Staff Scientist in the Fred Hutchinson Vaccine and Infectious Disease Division and an Affiliate Assistant Professor in the Departments of Global Health and Environmental and Occupational Health Sciences at the University of Washington.

An expert in Community-Based Participatory Research (CBPR), Historical Trauma and mixed methods research, Dr. Andrasik leads a robust Social and Behavioral Sciences research agenda for the HVTN. For nearly a decade, Dr. Andrasik has led the development and revision of the Behavioral Risk Assessment (BRA) utilized by the HVTN. Her efforts have focused on ensuring optimal behavioral and social factor risk reduction and assessment. In 2012, Dr.

Andrasik led the development of a social and behavioral sciences measures inventory across the HVTN. This effort facilitated the integration of social and behavioral sciences research (SBSR) across the Network and led to streamlining, harmonization and improvements in the quality of social and behavioral assessments. Following the 2012 development of the SBSR measures inventory, Dr. Andrasik has led ongoing efforts to identify risk criteria and optimize behavioral risk assessment. She has formed small protocol-specific working groups focusing on identifying region- and population-specific risk criteria to ensure optimal recruitment and assessment efforts. Small working groups have also worked to revise Phase 1, Phase II and efficacy trial behavioral risk assessments. Revisions are informed by analysis of behavioral data collected in HVTN phase 1, 2, 2b, and 2b/3 clinical trials as well as data obtained in the extant literature. Development and inclusion of assessment questions focuses on identifying variables that are most predictive of HIV risk and ensuring that assessments provide data that will be utilized and do not place unnecessary burden on the participants and site staff in the trials.

Dr. Andrasik has also participated in team efforts to analyze BRA data for inclusion in primary and secondary protocol manuscripts. More recently, she has engaged in an intensive systematic review to identify variables predictive of HIV seroconversion among sub- Saharan African heterosexual women. The manuscript will be submitted in the first quarter of 2020. In early 2020 another systematic review will be undertaken to identify variables predictive of HIV seroconversion among men who have sex with men in North and South America.

Chantel Smith

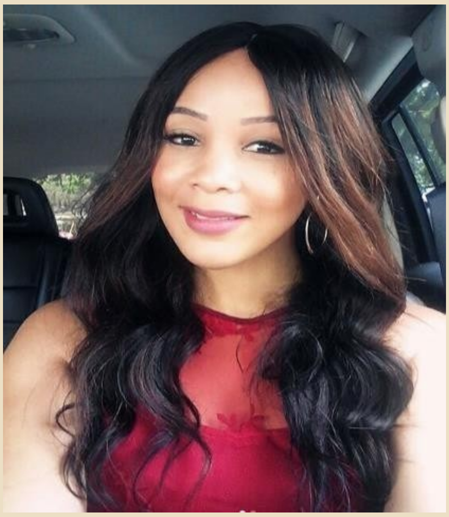

Dr Chantal A. Smith is the Technical Lead for Child and Adolescent Health Programmes at MatCH Institute, which is an indigenous South African public benefit organization that supports large scale HIV and ART services in a high burden, low- resourced environment.

As a Technical Lead, Dr Chantal Smith has focused on implementing health system strengthening strategies which integrate vertical programmes through the application of quality improvement principles at both health facility and health system levels.

She provides technical leadership and support to the South African Department of Health (national, provincial and district levels), in the areas of programming for children and adolescents living with HIV, adolescent pregnant women, non-infected adolescents and youth through a continuum of implemented strategies aimed at prevention and care.

One of her key activities that she has provided guidance on within the past 12 months, has been the implementation of Pre- Exposure Prophylaxis (PrEP) for high-risk HIV negative adolescent girls and young women (AGYW), aged 15-24 years.

Through this project, she has developed innovative screening tools and has provided technical leadership on the development of an integrated HIV prevention and sexual and reproductive health (SRH) package of care for AGYW.

Through various projects, she collaborates with a spectrum of stakeholders, from multi-national, national, provincial and district government, to international donors and local stakeholders such as managers from health facilities and civil society. In addition, she has provided strategic support to the Ministries of Health in both Ghana and Tanzania in the design, implementation and scale up of innovative paediatric, adolescent and youth-focussed interventions that were aimed at strengthening the existing components of the HIV prevention and treatment programme.

**Martina Morris**

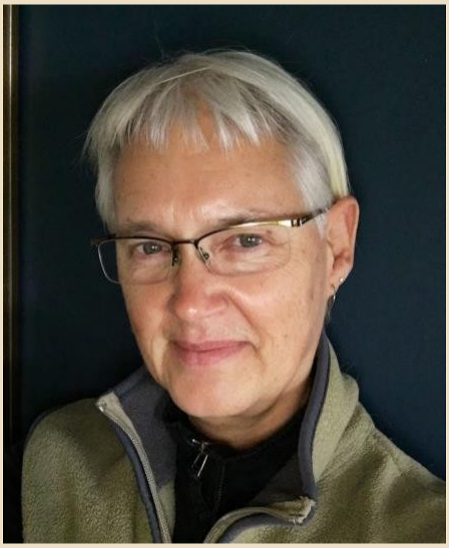

Martina Morris is an emeritus professor of Sociology and Statistics at the University of Washington. She has worked on the development of study designs and statistical methodology for social network data for over three decades, with specific applications to the projection and optimal control of HIV and other STIs. Her experience includes studies conducted in HIC and LMIC settings, with qualitative and quantitative components, and administration ranging from interviewer-based paper or mobile platforms to online data capture, with projects ranging from basic research to implementation science. She heads a large interdisciplinary team of researchers committed to the development of accessible reproducible research tools. They collectively develop and maintain a suite of open-source software packages written in the R programming language for statistical network analysis (statnet) and the mathematical modeling of infections across networks (EpiModel), teach annual courses on the use of these tools, and are collaborating to develop a new interactive graphical user interface for network data collection online (Network Canvas).

Kathryn O'Connell

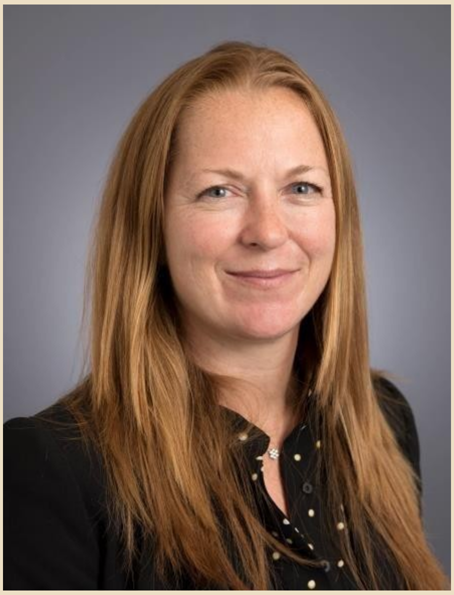

Kate O'Connell, PhD, MSc, MA, has more than 15 years' experience in monitoring and evaluation in public health research, including sexual and reproductive health and management of large-scale research projects. She has worked for the World Health Organization, Population Services International, ACTwatch and the World Bank and currently works for EngenderHealth, as the Director of Programme Impact, Research, and Evaluation. Kate has focused on the development and validation of several standardized questionnaires to address quality of life and health seeking behavior. Kate has authored more than 40 peer-reviewed publications addressing sexual and reproductive health, malaria, and health related quality of life.

Kate has lived across Asia and Africa and is currently based in Kampala, Uganda.

#### Meet theOrganizers

**Joseph Tucker**

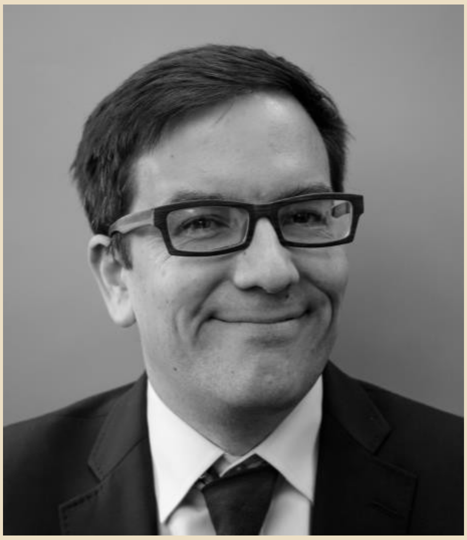

Joseph D. Tucker is an infectious diseases physician witha special interest in using crowdsourcing challenge contests to improve sexual health. He is an Associate Professor at the London School of Hygiene and Tropical Medicine and at the University of North Carolina at Chapel Hill School of Medicine. His team's ongoing research investigates challenge
contests to promote HIV, syphilis, HCV, and HBV testing. He is the Chairman of the Steering Committee of Social Entrepreneurship to Spur Health (SESH), a group focused on using crowdsourcing challenge contests to improve health. He has organized health hackathons in China and Nigeria. He has contributed to several WHO guidelines and serves as a member of the TDR Global Working Group. Joe received his BA from Swarthmore, MD from UNC, AM (RSEA) from Harvard, and PhD from the London School of Hygiene and Tropical Medicine.

Eneyi Kpokiri

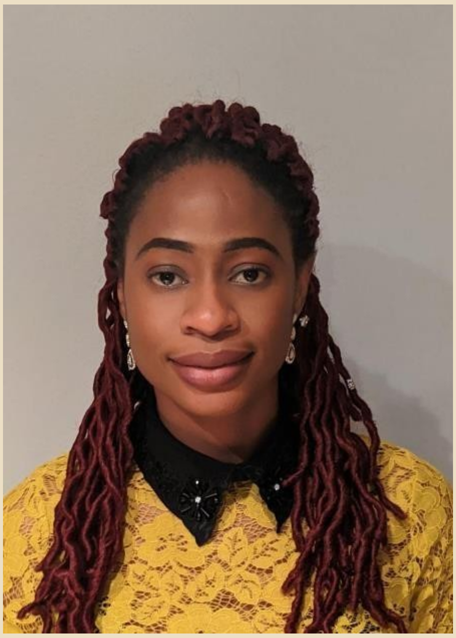

Eneyi Kpokiri, PhD is a Research Fellow in Social Innovation in Health at the London School of Hygiene and Tropical Medicine. She has conducted several innovation challenge contests in global health topics including AMR and access to diagnostics in LMICs. Her doctoral research from University College London, School of Pharmacy focused on improving the use of antibiotics by identifying strategies to support the implementation of effective antimicrobial stewardship programmes in low and middle-income hospital settings. She has experience in health services research using participatory and qualitative methods in low income settings. Her research is on exploring current public health services and practice patterns, identifying challenges and potential opportunities.

Megan Srinivas

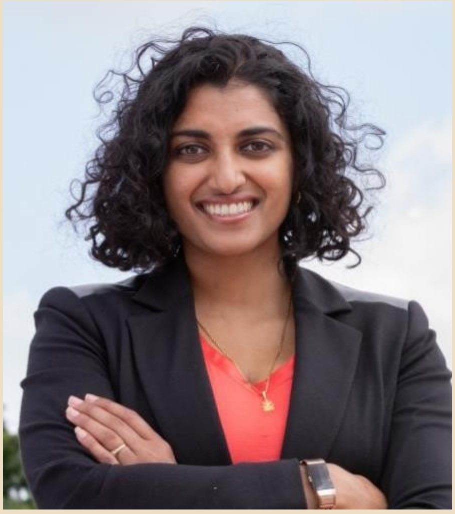

Dr. Megan Srinivas is an infectious disease fellow at the University of North Carolina. Her research focuses on how political change impacts access to reproductive health care, particularly in regards to the spread of STIs and HIV in rural areas. She worked for the World Food Prize Foundation in Kenya analyzing factors influencing household food security and was awarded the John Chrystal Award for outstanding contribution to hunger issues. In college, Dr. Srinivas co-founded Boston's Peer Health Exchange, a non-profit that teaches comprehensive sexual health education in socioeconomically-disadvantaged schools. For her senior thesis, she studied the evolution of malarial drug resistance in South America, changing national treatment policy in Peru and earning Harvard's Thomas Temple Hoopes Prize. During her Masters in Public Health, she investigated healthcare stigma/discrimination impeding HIV treatment in Brazil. Megan currently works with Project Echo to provide hepatitis C care via telehealth in the rural US. She is a national delegate to the American Medical Association and on the Infectious Disease Society of America Public Health Advisory Committee. Megan graduated Harvard College in 2009 with an AB cum laude in Human Evolutionary Biology and minors in Spanish, health policy, and Latin American studies.

She earned her Medical Degree from the University of Iowa in 2014, her MPH from Harvard in 2014, and completed her internal medicine residency at Johns Hopkins School of Medicine in 2017.

Juliana Anderson

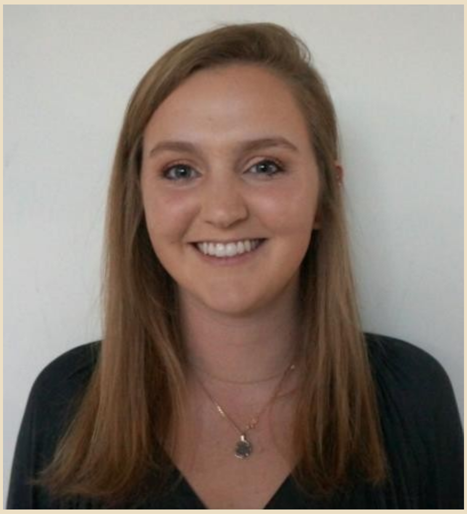

Juliana Anderson is an undergraduate at the University of North Carolina at Chapel Hill completing her B.A. in Chemistry and minors in Biology and Spanish for the Medical Professions.

Juliana has spent the last 7 months interning at the London School of Hygiene and Tropical Medicine (LSHTM) for Dr. Joseph Tucker. She has focused her time at LSHTM organizing the HRP hackathon, analyzing and scoring submissions, formatting and categorizing surveys, and drafting hackathon documents.

Dan Wu

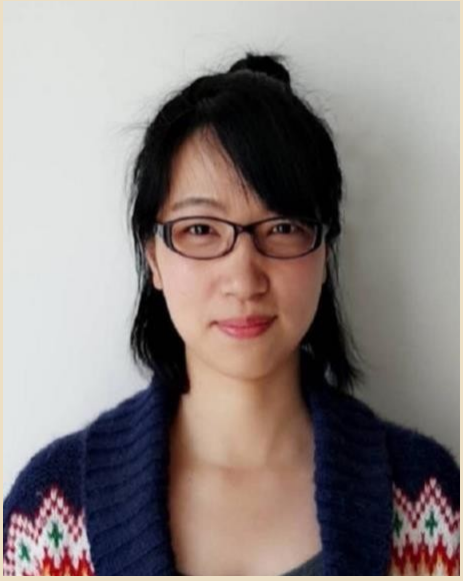

Dan Wu is a Newton International Fellow at The Academy of Medical Sciences and a research fellow at London School of Hygiene and Tropical Medicine, UK. She has a special research interest in understanding sexual health behaviors among key populations using both qualitative and quantitative methods. She has rich experiences of analyzing interview data and led several publications on studies using the mixed-methods approach. She has been intensively engaged in designing and managing projects using innovative strategies to improve sexual health services among marginalized populations in China.

Lianne Gonsalves

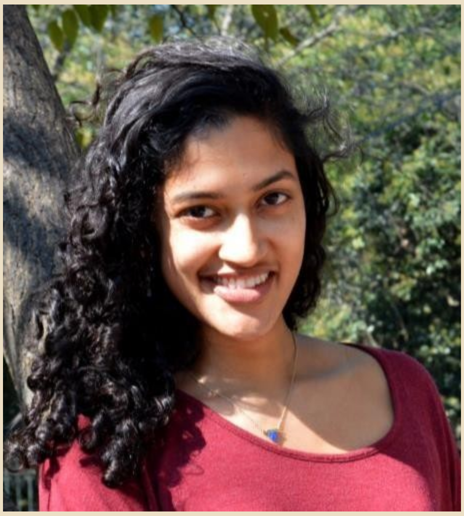

Lianne Gonsalves is a Technical Officer and has been with the WHO Department of Reproductive Health and Research since 2013. She is the Department's focal person for sexual health. In 2017, she led the development of WHO's *operational framework for sexual health and its linkages to reproductive health*.

Ongoing research involves exploring linkages between sexual functioning and other health conditions, as well as identifying global data and guidance needs related to sexual health and associated practices and behaviors. Previous experience includes working on sexual health and rights promotion among young people in Latin America; and integrated population, health, and environment programming in Ethiopia. She earned her MSPH at the Johns Hopkins Bloomberg School of Public Health, and is currently a PhD candidate at the University of Basel, Switzerland.

Meggie Mwoka

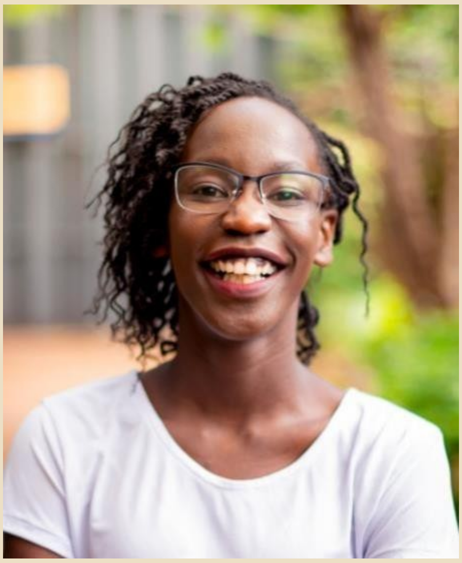

Dr. Meggie Mwoka is a medically qualified, global health specialist with 6 years' experience working in the NGO sector. Her public health and leadership experience span 10 countries, multiple projects on health systems strengthening and leading international teams. She currently works as a policy and research officer at the African Population and Health Research Center (APHRC) in a multi-country research, advocacy and capacity-strengthening project focused on addressing evident policy, institutional, and cultural dimensions of social exclusion that directly affect young people, women, and sexual minorities' access to sexual and reproductive health and rights. Her role involves stakeholder engagement, policy analysis, research and training on evidence-informed policymaking. She has experience conceptualizing and developing organizational strategies and coordinating and advising major national, regional and international programs and events in the realm of global/public health such as Africa Health Agenda International Conference, Medical Women's International Association Conference, Youth World Health Assembly. She is passionate about strengthening the African voice in the global health space and has delved into writing opinion pieces and blogs which have been published in key media channels such as El Pais and International Health Politics newsletter. Dr. Mwoka enjoys working with individuals from different backgrounds and sectors to share ideas, build networks and co-create solutions that will impact the community. She was selected as an UNLEASH 2017 SDG talent and has been awarded the Distinguished leadership among young women doctors by the African Medical Women International Association.

Evelyn Gitau

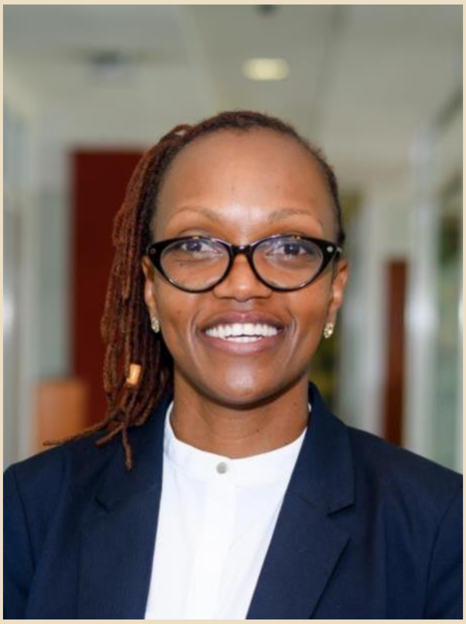

Dr. Evelyn Gitau is the Director of Research Capacity Strengthening division at the African Population and Health Research Center (APHRC) where her main role is to expand opportunities for African scholars to become research leaders and to support the growth of APHRCs signature fellowship program, Consortia for Advanced Training in Africa (CARTA). Evelyn was previously a Programme Manager at the African Academy of Sciences where she was responsible for the Grand Challenges Africa.
She has over 16 years' experience in medical research and sits on the advisory board of several organisations including the Independent Scientific Advisory Board (ISAB), Malawi-Liverpool- Wellcome Trust Clinical Research Programme College of Medicine, Blantyre, Malawi, University of Oxford (MSc International Health and Tropical Medicine), the Crick-Africa Network and the Investment Committee -Grand Challenges Canada

**Lisa Omondi**

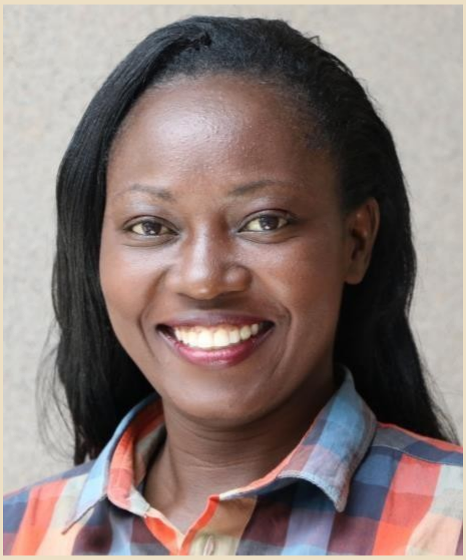

Lisa Omondi, is the lead Program Assistant in the Research Capacity Strengthening (RCS) Division at the African Population and Health Research Center Kenya. She is in charge of a range of administrative and capacity building initiatives in the RCS Division, as well as coordinating routine Brown bag scientific presentations at APHRC. Lisa also provides support to the African Doctoral Dissertation Research Fellowship (ADDRF) program.

Prior to joining APHRC, Lisa worked as a Training Assistant at ICIPE. She possess over 10 years' experience in programming focused on Training and scientific capacity building. She hold a Bachelor's degree in Human Resource Management from Mount Kenya University, Kenya.
Lisa is driven by philanthropy work. In her spare time, she participates in philanthropy activities such as visiting and assisting needy children.

#### Steering Committee

We are grateful for the support and wisdom of our steering committee. The following individuals joined our steering committee and met by teleconference to discuss progress and make key decisions: Lianne Gonsalves (Co-Chair, World Health Organization, WHO); Joseph Tucker (University of North Carolina and the London School of Hygiene and Tropical Medicine, Co-Chair); Lale Say (WHO); Megan Srinivas (UNC); Nathalie Bajos – French National Institute of Health and Medical Research (INSERM); Emma Slaymaker (LSHTM; Annette Sohn (TREAT Asia/The Foundation for AIDS Research, amfAR); Laura Lindberg (Guttmacher Institute); Pedro Nobre (World Association for Sexual Health); Linda-Gail Bekker (University of Cape Town/International Aids Society); Cesar Carcamo (Universidad Peruana Cayetano Heredia); Eneyi Kpokiri (SESH Global); Kaye Wellings (LSHTM); Boniface Ushie (African Population Health Research Center).

**Supplementary file 4: Survey questionnaire for Delphi**

#### HRP Delphi Survey

Thank you for taking part in this survey. This will take about 20 minutes and will help us to synthesize ideas for the HRP sexual and reproductive health hackathon.

3. **Name:**

First Name Last Name

#### Consensus Statement

The next section has a draft consensus statement that has been revised based on the initial Delphi survey.

A sexual health survey instrument should do the following:

**Design Stage:**

| | Strongly agree | Agree | Neutral | Disagree | Strongly disagree |
|---|---|---|---|---|---|
| 4. Be inclusive of the local population’s needs, desires, and preferences | ✘ | ✘ | ✘ | ✘ | ✘ |
| 5. Promote sexual and reproductive health and positive choices and agency instead of absence of disease or poor outcomes | ✘ | ✘ | ✘ | ✘ | ✘ |
| 6. Give agency to individuals to make their own decisions about sexual and reproductive health | ✘ | ✘ | ✘ | ✘ | ✘ |
| 7. Promote well-being and reflect the lived reality of the user journey (the experience of taking the survey in the local context) | ✘ | ✘ | ✘ | ✘ | ✘ |
| 8. Ensure that the survey and its design, implementation, and dissemination are directly informed by local people | ✘ | ✘ | ✘ | ✘ | ✘ |
| 9. The survey items focus on sexual well-being rather than disease. | ✘ | ✘ | ✘ | ✘ | ✘ |
| 10. Acknowledges the broader determinants of sexual health outcomes | ✘ | ✘ | ✘ | ✘ | ✘ |
| 11. Appreciate the local and national norms that are related to age of consent, homosexuality, abortion, gender issues, and related macro-level factors | ✘ | ✘ | ✘ | ✘ | ✘ |

12. **Comments on any of the 'design stage' statements above**

**Training:**

| | Strongly agree | Agree | Neutral | Disagree | Strongly disagree |
|---|---|---|---|---|---|
| 13. **Ensure that organizers are trained in identification of** violence in sexual and reproductive health services and assist in linking those affected to appropriate services | ✘ | ✘ | ✘ | ✘ | ✘ |
| 14. **Includes female interviewers, gender-specific topics, recognizes the importance of gender throughout the research project.** | ✘ | ✘ | ✘ | ✘ | ✘ |

15. **Comments on any of the 'training' statements above.**

**Implementation:**

| | Strongly agree | Agree | Neutral | Disagree | Strongly disagree |
|---|---|---|---|---|---|
| 3. **Ensure the anonymity and privacy of survey** participants | ✗ | ✗ | ✗ | ✗ | ✗ |
| 4. **Promote, protect, and fulfill human rights, including sexual rights** | ✘ | ✘ | ✘ | ✘ | ✘ |
| 5. **Take a broad bio-psycho-social perspective on SRH when adapting survey items for the local context** | ✘ | ✘ | ✘ | ✘ | ✘ |

6. **Comments on any of the 'implementation' statements above.**

**7. Other comments or issues that are not well covered above?**

### Revised Survey Domains

Please review each of the items listed below carefully and assign one of the following levels. Please very cautiously select what items are categorized as tier 1.

**Tier 1**: Highest priority items that should be included in a 10-minute survey instrument

**Tier 2**: Medium priority items that should be included in a 20-minute survey instrument

**Tier 3**: Lowest priority items to be included in a 30-minute survey instrument.

#### Sexual health outcomes (HIV/STIs and reproductive outcomes)

**Sexual health outcomes (HIV/STIs and reproductive outcomes)**

| | Tier 1 | Tier 2 | Tier 3 |
|---|---|---|---|
| **8. Number of pregnancies (including unintended/intended pregnancies)** | ✘ | ✘ | ✘ |
| **9. Number of spontaneous abortions(miscarriage)** | ✘ | ✘ | ✘ |
| **10. Number of abortions** | ✘ | ✘ | ✘ |
| **11. Contraceptive practices (including fertility intention (infertility), utilization of contraceptives, and types of contraceptives)** | ✘ | ✘ | ✘ |
| **12. STIs history** | ✘ | ✘ | ✘ |
| **13. HIV status** | ✘ | ✘ | ✘ |

**Other sexual health outcome considerations:**

### Sexual Practices/Sexual Satisfaction

#### Sexual practices

| | Tier 1 | Tier 2 | Tier 3 |
|---|---|---|---|
| 25. Type of sexual intercourse | ✘ | ✘ | ✘ |
| 26. Masturbation | ✘ | ✘ | ✘ |
| 27. Sexual frequency | ✘ | ✘ | ✘ |
| 28. Sex toys | ✘ | ✘ | ✘ |
| 29. Pornography | ✘ | ✘ | ✘ |
| 30. Group sex | ✘ | ✘ | ✘ |
| 31. Drug/alcohol related sexual practices | ✘ | ✘ | ✘ |

#### Sexual satisfaction, well-being, and (dys)function

| | Tier 1 | Tier 2 | Tier 3 |
|---|---|---|---|
| 1. Level of satisfaction | ✘ | ✘ | ✘ |
| 2. Causes of dissatisfaction | ✘ | ✘ | ✘ |
| 3. Perceived discrepancy between own and partner's: sexual satisfaction, sexual desire , and sexual self-esteem | ✘ | ✘ | ✘ |
| 4. Sexual well-being | ✘ | ✘ | ✘ |
| 5. Feelings of safety and security (safety with sexual intercourse and within relationships) | ✘ | ✘ | ✘ |
| 6. Sexual (dys)function (including problems surrounding sexual function and medicines/methods to promote sexual function) | ✘ | ✘ | ✘ |

**Other sexual practices/sexual satisfaction considerations:**

### Social Norms/Sexual Rights

##### Perceptions related to sex/sexuality

| | Tier 1 | Tier 2 | Tier 3 |
|---|---|---|---|
| 38. Homosexuality | ✘ | ✘ | ✘ |
| 39. Minimum age of acceptable marriage (female and male) | ✘ | ✘ | ✘ |
| 40. Sexual violence/non-consensual sex | ✘ | ✘ | ✘ |
| 41. Abortion | ✘ | ✘ | ✘ |
| 42. Contraception | ✘ | ✘ | ✘ |
| 43. Multiple concurrent partners (male and female) | ✘ | ✘ | ✘ |
| 44. Sexual drives (male and female) | ✘ | ✘ | ✘ |
| 45. Sexual satisfaction and wellbeing (male and female) | ✘ | ✘ | ✘ |
| 46. Gender equality related sexuality | ✘ | ✘ | ✘ |

##### Sexual rights (availability of health services/access to healthcare)

| | Tier 1 | Tier 2 | Tier 3 |
|---|---|---|---|
| 47. Contraception | ✘ | ✘ | ✘ |
| 48. Abortion | ✘ | ✘ | ✘ |
| 49. Intimate partner violence including sexual violence | ✘ | ✘ | ✘ |
| 50. Consented sex | ✘ | ✘ | ✘ |
| 51. LGBTQ rights | ✘ | ✘ | ✘ |
| 52. Power dynamics | ✘ | ✘ | ✘ |

##### Non-sexual gender roles and expectations

| | Tier 1 | Tier 2 | Tier 3 |
|---|---|---|---|
| 53. Gender role and expectations (non-sexual) | ✘ | ✘ | ✘ |

##### Other social norms/sexual rights considerations:

**Sexualbiography(i.e.sexualhistory)**

| | Tier 1 | Tier 2 | Tier 3 |
|---|---|---|---|
| • Gender identity | ✗ | ✗ | ✗ |
| • Sexual orientation | ✗ | ✗ | ✗ |
| • Number of male, female, and other partners | ✗ | ✗ | ✗ |
| • Number ofnon-penetrative partners | ✗ | ✗ | ✗ |
| • Number of partners met through internet | ✗ | ✗ | ✗ |
| • Firstpartner(includingage,consensualornon-consensual,circumstances surrounding first sex) | ✗ | ✗ | ✗ |
| • Current/lastpartner(includingconsensualornon-consensual,meansforfindingthe sexual partner, and circumstances surrounding last sex) | ✗ | ✗ | ✗ |
| 61. Transactional sex | ✗ | ✗ | ✗ |
| 62. Non-consensual sex | ✗ | ✗ | ✗ |
| 63. Sexual violence | ✗ | ✗ | ✗ |
| 64. Partner history | ✗ | ✗ | ✗ |
| 65. Trend of steady partners or marriages | ✗ | ✗ | ✗ |
| 66. New partners | ✗ | ✗ | ✗ |
| 67. Concurrent relationships | ✗ | ✗ | ✗ |

**Other sexual biography considerations:**

#### Socio-demographic Information

| | Tier 1 | Tier 2 | Tier 3 |
|---|---|---|---|
| 68. Respondent ID | ✘ | ✘ | ✘ |
| 69. Biological sex | ✘ | ✘ | ✘ |
| 70. Gender identity | ✘ | ✘ | ✘ |
| 71. Age | ✘ | ✘ | ✘ |
| 72. Race& ethnicity | ✘ | ✘ | ✘ |
| 73. Marital status / cohabitation status | ✘ | ✘ | ✘ |
| 74. Number of household members | ✘ | ✘ | ✘ |
| 75. Relationship history | ✘ | ✘ | ✘ |
| 76. Education | ✘ | ✘ | ✘ |
| 77. Highest level of formal education | ✘ | ✘ | ✘ |
| 78. Children | ✘ | ✘ | ✘ |
| 79. Financial resources / employment status | ✘ | ✘ | ✘ |
| 80. Financial independence | ✘ | ✘ | ✘ |
| 81. Monthly household income/weighted financial system | ✘ | ✘ | ✘ |
| 82. Type of (regular) resources? | ✘ | ✘ | ✘ |
| 83. Rural/urban | ✘ | ✘ | ✘ |
| 84. Disabilities | ✘ | ✘ | ✘ |

**Draft Survey Instrument**

**General Introduction**
[For the survey organizers]
**Process:**
The following instrument was generated via a multi-step process that started with the survey instrument created during a 3-day sexual health hackathon held in Nairobi, Kenya in January 2020. A hackathon is a sprint-like event that brings together individuals with diverse backgrounds to solve a problem.[1] A hackathon can tap into participants' experiences and expertise to generate high-quality outputs in a transparent and systematic way.[2] The resulting survey was discussed in the final session with all hackathon participants. The UNDP/UNFPA/UNICEF/WHO/World Bank Special Programme of Research, Development and Research Training in Human Reproduction's (HRP) hackathon organizing committee used comments from this final group session to revise the survey. Each individual who attended the hackathon received a copy of the revised survey and was given the opportunity to provide written feedback and suggestions. The organizing committee reviewed these comments, and after thorough discussion, completed the third revision of the survey. The two lead facilitators of the hackathon did a further round of revisions. Their suggestions and edits were forwarded back to the organizing committee and steering committees for review and feedback. This overall process was repeated once more, and the document incorporates all feedback to date.

This survey is intended for population-based research studies or surveillance in public health, but may be useful for epidemiological analyses, clinical trials, or other types of study. The items below are considered as a core set of questions. This core set can be implemented as part of a larger established population-based survey. These questions include addressing some sensitive issues (for example, related to abortion, same-sex behaviours, and sexual violence), and are amenable to adaptation, which may require more extensive field testing in the local context, or in some settings, may need to be omitted.

For more information about implementation considerations, see our consensus statement.

**[For the participant, before the survey]**
This [section of the] survey is about sexual and reproductive health experiences. This information will be used to inform health policy, improve health care and health outcomes. This survey is designed to be completed by a wide range of people and so some questions may not apply to you. Some of the questions may surprise you, may cause embarrassment, and/or may be difficult to answer. Please remember you can choose not to answer any question you do not want to. All your responses will be completely confidential and kept anonymous.[3] We thank you for your participation.

**Survey Items**

**A: Socio-demographics & health**

[A1] What sex are you on your birth certificate *(pending location, replace with context-specific ID document assigned at birth)*?

a. Male
b. Female

[A2] How old were you at your last birthday? XXX years or Don't know (as locally appropriate)

[A3] How many times have you been married or lived with a partner (co-habited)?

---

[1] *Health Hackathon Handbook – MIT Hacking Medicine. 2016. http://hackingmedicine.mit.edu/healthcare-hackathon-handbook/*

[2] *Tucker JD, Tang W, Li H, et al. Crowdsourcing designathon: a new model for multisectoral collaboration. BMJ Innovations 2018;**4:**46-50.*

[3] *If supported by your ethics application. Will need to specify at time of application and amend this language pending result.*

a. ____ times
b. I have never been married or lived with a partner

*[skip to A5 if answer to A3 is b.]*

[A4] How old were you when you first started living with a partner or spouse? XXX years or Don't know (as locally appropriate)

[A5] Are you at present…

a. Single
b. Married, living with spouse
c. Married, not living with spouse
d. Divorced
e. Widowed

[A6] Now thinking about your health currently, how is your health in general?

a. Very good
b. Good
c. Fair
d. Poor
e. Very Poor

[A7, for field testing] Do you currently have any mental or physical illness or disability that affects you in your everyday life? By affecting your life, we mean limiting your usual activities in any way.

a. Yes. If yes, please list the illnesses and/or disabilities.
b. No

**B. Sexual health outcomes**

The next section asks about pregnancy and other sexual health outcomes.

[B1] [to women] To the best of your knowledge, how many times have you been pregnant to date? XXX or don't know
[B1] [to men] To the best of your knowledge, how many times have you gotten a woman pregnant to date? XXX or don't know

[B2] *only ask women who reported 1 or more pregnancies at B1:* How old were you at the time of your **first** pregnancy (including any pregnancies that did not result in a live birth)? (age at the end of the pregnancy) XXX years or Don't know (as locally appropriate)
*For men* How old were you the **first** time you got a woman pregnant (including any pregnancies that did not result in a live birth)? (age at the end of the pregnancy) XXX years or Don't know (as locally appropriate)

[B3] *only ask men & women who reported 2 or more pregnancies at B2:* How old were you at the time of your **last** pregnancy (including any pregnancies that did not result in a live birth)? (age at the end of the pregnancy) XXX years or Don't know (as locally appropriate)
*For men* How old were you the **last** time you got a woman pregnant (including any pregnancies that did not result in a live birth)? (age at the end of the pregnancy) XXX years or Don't know (as locally appropriate)

[B4, for field testing] *For women with last pregnancy in last 5 years (B3):* When you became pregnant with your last (or current) pregnancy, how much did you personally want to become pregnant at that time?

a. Did not want **at all** to become pregnant at that time
b. Somewhat did **not want** to become pregnant at that time
c. Unsure about becoming pregnant at that time
d. Somewhat **wanted** to become pregnant at that time
e. Wanted **very much** to become pregnant at that time

[B5, for field testing] *if male & last pregnancy in last 5 yrs (B3)*: When your partner became pregnant with their last pregnancy, how much did you personally want your partner to become pregnant at that time?

a. Did not want **at all** to get partner pregnant at that time
b. Somewhat did **not want** to get partner pregnant at that time
c. Unsure about getting partner pregnant at that time
d. Somewhat **wanted** to get partner pregnant at that time
e. Wanted **very much** to get partner pregnant at that time

[B6] If reported 1 or more pregnancies at B1: How many of these pregnancies resulted in:
- Live birth (baby born alive) *<enter number; include 'don't know' response option – not just for men>*
- Abortion (medical or surgical for any reason*; include 'don't know' response option – not just for men*)
- Miscarriage at:
    - < 12 weeks pregnancy *<enter number; include 'don't know' response option – not just for men >*
    - More than 12 weeks pregnancy *<enter number; include 'don't know' response option – not just for men >*
    - How many required an additional medication of procedure? *<enter number; include 'don't know' response option – not just for men >*
- Still birth or baby born without heartbeat/not breathing *<enter number; include 'don't know' response option – not just for men >*
- Currently pregnant

[B7] *only ask men & women who reported 1 or more live/still birth at B2:* How old were you when you/your partner **first** gave birth? XXXX years or Don't know (as locally appropriate)

[B8, field testing] Have you ever had a time lasting 1 year or longer when you and your partner were trying to get pregnant and it did not happen?

a. Yes
b. No

These following questions ask about the human immunodeficiency virus (HIV, the virus that causes AIDS), and sexually transmitted infections (STIs).

[B9, for field testing] When, if ever, were you last **tested** for HIV?
a. In the last year
b. More than 1 year ago
c. Never
d. Don't know

[B10, for field testing, assuming no mandatory reporting requirements in country] If tested: What was the result of your last HIV test?
a. I have HIV
b. I do not have HIV
c. I am still waiting for the test results
d. I don't know

e. I prefer not to say

[B11] **Aside from HIV**, when, if ever, were you last **tested** for sexually transmitted infections (STIs) (e.g. gonorrhoea, chlamydia, syphilis, herpes, trichomoniasis)?

a. In the last year
b. More than 1 year ago
c. Never
d. Don't know

[B12] **Aside from HIV**, when, if ever, did you last receive **treatment** for a STI (e.g. gonorrhoea, chlamydia, syphilis, herpes, trichomoniasis) (either self-treatment or treatment from a doctor)?
List month and year (XX/XX) or never. If unclear, mark last year / more than a year ago / don't know

Non-consensual / violence (sex against your will). The next question is about non-consensual sexual situations that anyone may have encountered. We understand that these are sometimes difficult to think/talk about, and you can skip any questions you feel uncomfortable answering. (This section needs to field tested)

[B13, field testing] Currently, in your everyday life (i.e., at work, on the street, at home), how vulnerable do you feel to sexual assault? 5-point Likert response scale:
1 – not at all safe
2 – somewhat unsafe
3 – neither safe or unsafe
4 – somewhat safe
5 – signifying completely safe

[B14A, field testing] Have you ever been either forced or frightened by another person into doing something sexually that you did not want to do? *<response options:*

a. Yes
b. Can't remember
c. This has not happened to me

*If 'this has not happened to me', then proceed to question C1.*

[B14B, field testing] Has this happened to you more than once? *<response options: Yes/ No/ >*

*If yes to B14B, then ask B15A/B. If no, skip B15.(field testing)*
[B15A] How old were you the **first** time this happened? *<response options: XXX years /* Don't know (as locally appropriate)
[B15B] How old were you the **last** time this happened? *<response options: XXX years /* or Don't know (as locally appropriate)

**C. Sexual biography**

The next section asks questions about your sexual experience. By 'sexual experience' we mean any kind of contact with another person that you felt was sexual. It could be kissing, touching, intercourse, or any other form of sex.

[C1] Which of these statements best describes you? (please choose all that apply)

a. I have had sexual experiences only with males, never with females
b. I have had sexual experiences mostly with males, and at least once with a female
c. I have had sexual experiences both with males and females
d. I have had sexual experiences mostly with females, and at least once with a male
e. I have had sexual experiences only with females, never with males

f. I have (also) had sexual experience with individual(s) who do not identify as male or female
g. I have not had any sexual experience
h. I do not want to answer

[C2] How old were you the first time you had sex with someone? That is, had any sexual contact involving the genital area, including oral sex, vaginal sex, anal sex, or [*insert culturally appropriate terms].* Please type in the age in years. Please estimate the age if you can't say exactly**.** XXXX years or Don't know

[C3] [*ASK ONLY IF REPORTED SAME-SEX EXPERIENCE].* The first time you had sex, was this person (select all that apply)…

a. Male
b. Female
c. Other
*if other please specify*

[C4] How old was your partner at the time you first had sex? XXXX years or Don't know
Please estimate if you do not know exactly.

- If respondent unsure, then ask: Was your partner older than you, younger than you, or about the same age as you?
- If older or younger, ask: By how many years?
    a. 1-2 years
    b. 3-5 years
    c. 6-10 years
    d. 10+ years

[C5, for field testing] Which statement applies best to the first time you had sex? **(choose all that apply)**

a. I wanted it
b. I was forced into doing it
c. I forced the other person
d. Can't remember

[C6] *If answered a-f for C1 or if C1 was not asked, then ask:* What precautions against pregnancy or HIV/STIs did you use the first time you had sex, if any? (choose all that apply)

a. No precautions
b. Male Condom
c. Female Condom
d. Birth control/Oral contraceptive pill
e. Morning after pill/Emergency oral contraceptive pill
f. IUD/Coil/Loop
g. Emergency intrauterine device (IUD)/Coil/Loop
h. Cap/Diaphragm
i. Injections
j. Spermicides (foams/gels/sprays/pessaries)
k. He/I withdrew
l. Made sure it was safe time period in my/her monthly cycle (calendar method/safe period)
m. Partner was/I had been sterilized
n. Other method of protection (please say what)
o. Don't know

[C7] In your life so far, how many people have you had sex with? [stratified by gender if reported same-sex experience at C1 – even if hadn't had actual same-sex **sex**] That is any sexual contact involving the genital area, including oral sex, vaginal sex, anal sex, or [*insert culturally appropriate terms].* Please include everyone you have ever had sex with, whether it was just once or multiple times, with a stranger, regular partner, or husband/wife.

[C8] In the last year, how many people have you had sex with? That is any sexual contact involving the genital area, including oral sex, vaginal sex, anal sex, or [*insert culturally appropriate terms]*? [stratified by gender if reported same-sex experience at C1 or C3 – even if hadn't had actual same-sex **sex** – or if ask about number of same-sex partners in lifetime [C7]and report 1+ ever]

[C9] In the last 4 weeks**,** how many people did you have sex with? That is any sexual contact involving the genital area, including oral sex, vaginal sex, anal sex, or [*insert culturally appropriate terms]*? [stratified by gender if reported same-sex experience at C1 or C3 – even if hadn't had actual same-sex **sex** – or if ask about number of same-sex partners in the last year [C8] and report 1+ ever]

<u>Transactional sex (both/either way).</u> The next section is about situations when sex is exchanged for goods, services, or money.

[C10] When, if ever, was the last time you **gave** money, material goods, favours, gifts, drugs, or shelter in exchange for sex? By material goods, we mean things like food, rent, clothes/shoes/cell phones, cosmetics, transport, good marks in school or school fees, or items for your partner or their children, their family, or themself.

a. In the last year
b. More than a year ago
c. Never

[C11A] When, if ever, was the last time you **received** money, material goods, favours, gifts, drugs, or shelter in exchange for sex? By material goods, we mean things like food, rent, clothes/shoes/cell phones, cosmetics, transport, good marks in school or school fees, or items for you or your children, your family, or yourself.

a. In the last year
b. More than a year ago
c. Never

**D. Sexual Practices** (will field test questions on a type of sexual act)

[D1] ONLY ASK IF HAD SEX IN THE LAST YEAR (C8 ≥1): How many times have you had sex with another person or people in the last 4 weeks? That is any sexual contact involving the genital area, so including oral sex, vaginal sex, anal sex, or [*insert culturally appropriate terms]*

[For anyone reporting have sex in the last year, (C8 ≥1)] <u>Questions D2-D9</u> are about the **last** time you had sex, meaning the most recent time you had **any** sexual contact with another person.

[D2] When did you last have sex? Mm/yy

[D3] Which one of these descriptions applies best to you and ***(that person)*** at the time you **most recently** had sex? Only give one answer

a. We were living together as a couple / married / in a civil partnership at the time
b. We were in a steady relationship at the time
c. We used to be in a steady relationship, but were not at that time
d. We had known each other for a while, but were not in a steady relationship
e. We had recently met
f. We had just met for the first time

[D4] How old was that person when you last had sex together? XXX years or Don't know
Please estimate if you do not know exactly.

- If respondent unsure, then ask: Was your partner/s older than you, younger than you, or about the same age as you?
- If older or younger, ask: By how many years?
    a. 1-2 years
    b. 3-5 years
    c. 6-10 years
    d. 10+ years

[D5] *If reported same-sex experience at B1*: What was your partner's sex:

[D6] To which of the ethnic groups do you consider this person belongs? [for field testing and localization]

[D7, for field testing] The **last** time you had sex with this person, which of the following did you do? (Mark all that apply)

a. You performed oral sex on them
b. They performed oral sex on you
c. You had penile-vaginal intercourse
d. You inserted something into their vagina/They inserted something into your vagina (includes fingers, hands, dildos, toys, or other sexual aids)
e. You had receptive penile-anal intercourse
f. You had insertive penile-anal intercourse
g. You inserted something in their anus (includes fingers, hands, dildos, toys, or other sexual aids)
h. They inserted something in your anus (includes fingers, hands, dildos, toys, or other sexual aids)
i. Other sexual contact not listed here

[D8] Which precautions against pregnancy or HIV/STIs did either of you take the last time you had sex together?

a. No precautions
b. Male Condom
c. Female Condom
d. Birth control/Oral contraceptive pill
e. Morning after pill/Emergency oral contraceptive pill
f. IUD/Coil/Loop
g. Emergency intrauterine device (IUD)/Coil/Loop
h. Cap/Diaphragm
i. Injections
j. Spermicides (foams/gels/sprays/pessaries)
k. He/I withdrew
l. Made sure it was safe time period in my/her monthly cycle (calendar method/safe period)
m. Partner has been /I have been sterilized
n. Other method of protection (please say what)
o. Don't know

[D9] How pleasurable did **you** find the last time you had sex?

a. very pleasurable
b. pleasurable
c. neutral
d. unpleasurable
e. very unpleasurable).

For the types of sex that did not happen during the last time that you had sex (per participant's answer to D7), the following questions will be asked:
[D10] When, if ever, was the last time you performed oral sex on someone, that is your mouth on their genital area?

a. In the last 4 weeks

b. more than 4 weeks ago, but within the last year
c. more than 1 year ago
d. never

[D11] When, if ever, was the last time someone performed oral sex on you? That is, their mouth on your genital area?

a. In the last 4 weeks
b. more than 4 weeks ago, but within the last year
c. more than 1 year ago
d. never

[D12] When, if ever, was the last time you had vaginal sex with someone (*woman/man*)? Vaginal sex is a penis in a vagina.

a. In the last 4 weeks
b. more than 4 weeks ago, but within the last year
c. more than 1 year ago
d. never

[D13] When, if ever, was the last time you were anally stimulated/you anally stimulated someone (*woman/man*)? Anal stimulation is hands, dildo or other sexual aids in the anus (rectum or back passage).

a. In the last 4 weeks
b. more than 4 weeks ago, but within the last year
c. more than 1 year ago
d. never

[D14A] When, if ever, was the last time you had **receptive** anal sex with someone (*woman/man*)? Receptive anal sex is having a penis inserted into your anus (rectum or back passage).

a. In the last 4 weeks
b. more than 4 weeks ago, but within the last year
c. more than 1 year ago
d. never

[D14B] When, if ever, was the last time you had **insertive** anal sex with someone (*woman/man*)? Insertive anal sex is inserting a penis into another person's anus (rectum or back passage).

a. In the last 4 weeks
b. more than 4 weeks ago, but within the last year
c. more than 1 year ago
d. never

[D15] When, if ever, was the last time you had manual sex (field test versus manual stimulation) (with a man/woman-) that is, a hand/hands or sexual aids (i.e., dildos, toys) on or in a genital area?

a. In the last 4 weeks
b. more than 4 weeks ago, but within the last year
c. more than 1 year ago never

[D16] Solo masturbation: When, if ever, did you **last** masturbate, that is, arouse and pleasure **yourself** sexually?

a. In the last 4 weeks
b. more than 4 weeks ago, but within the last year
c. more than 1 year ago
d. never

[D17] In general, how satisfied have you been with your sex life in the last year?

a. very satisfied
b. satisfied
c. neutral
d. dissatisfied
e. very dissatisfied

**E. Social Perceptions/Beliefs** (to field test)

For E1 – E13, please read the following statements and say whether **you**:

1. Strongly agree
2. Agree
3. Disagree
4. Strongly disagree
5. Prefer not to answer

[E1] Sex education promotes sexual activity among young people.

[E2] A woman has the right to say 'no' to sex if she does not want it.

[E3] A man has the right to say 'no' to sex if he does not want it.

[E4] It is acceptable for a woman to have sex before marriage.

[E5] It is acceptable for a man to have sex before marriage.

[E6] Having sex that is pleasurable is important for a woman's sex life and general well being

[E7] Having sex that is pleasurable is important for a man's sex life and general well being

[E8] Sex between two consenting adult women is always wrong.

[E9] Sex between two consenting adult men is always wrong.

[E10] Men naturally have more sexual needs than women.

[E11] It is okay for a woman to use a modern contraceptive method/family planning (*e.g.* birth control/oral contraceptive pills, injection, implants, loop or coil (IUD), condoms, etc) to avoid or delay pregnancy if she wishes.

[E12] It is okay for a woman to [have an abortion / terminate a pregnancy] if she does not want to have a child

[E13] Who do you think should decide whether a woman [has an abortion/ terminates a pregnancy]?

a. Mainly her decision
b. Mainly her husband's or partner's decision
c. They should decide together
d. Others (please specify)
e. No response

**F. Identity**

[F1] Do you think of yourself as…?

a. Male
b. Female
c. Another gender identity not listed here (please specify)

[F2] Do you think of yourself as ...

a. Heterosexual or straight
b. Gay, lesbian, or homosexual

c. Bisexual
d. Pansexual
d. Asexual
e. Not sure; undecided /another identity not listed here
f. Do not wish to answer

[F3] Education < locally determined >

[F4] Which best describes your employment? < locally determined >

[F5, for field testing] How often do you find your household not having enough resources to obtain what they need to live day to day?

a. Every day
b. At least once per week
c. At least once per month
d. At least once per year
e. Never

[F6] To which of the ethnic groups on this card do you consider you belong? < locally determined >

[F7A] What is your current religion?> < locally determined >

[F7B, for field testing] How religious do you consider yourself? (5-option Likert scale with 1 signifying not religious at all and 5 signifying very religious)

**Sexual rights**

Given the importance of sexual rights in a broad sexual health framework, this is an important topic.

**Discrimination against sexual minorities**
[ask only if F2 response is b-f]
[F8] Have you ever been discriminated against because of sexual orientation? (Yes/ No)

If yes, when was the last time you were discriminated against?

a. In the last year
b. More than 1 year ago
c. Don't know/Prefer not to answer

[F9] For the following 9 statements (A-I), please mark when, if ever, you have experienced any of the following on the grounds of your sexual orientation? <1. In the last year, 2. More than 1 year ago, 3. Never, 4. Don't know/prefer not to answer>

[A] I have been insulted or threatened.

[B] I have been beaten, pushed or kicked

[C] My belongings have been destroyed or damaged

[D] I was not given a job or was dismissed from my job

[E] I was treated in a discriminatory way by a healthcare professional

[F] I was denied medical treatment

[G] I was jailed, prosecuted or denied legal services

[H] I was asked to leave my home or thrown out of my accommodations

[I] I was forced to engage in a sexual act, sexually assaulted, or raped

Thank you for completing the survey. [THE END]

**Appendix 1: Organizers, Steering Committee, and Hackathon Facilitators/Participants (names listed in alphabetical order)**

**Organizers** – individuals who organized the project from conception to post-hackathon synthesis.

Juliana Anderson
Evelyn Gitau
Lianne Gonsalves
Eneyi Kpokiri
Meggie Mwoka
Lale Say
Megan Srinivas
Joe Tucker
Dan Wu

**Hackathon Facilitators** – facilitators each led one of the five discussion groups for survey construction (1. Implementation, 2. General Information, 3. Sexual Biography, 4. Sexual Practices, 5. Sexual Norms and Understanding). Co-lead facilitators advised on the overall construction of the survey and advised each of the groups throughout the hackathon.

Noor Ani Ahmad
Nathalie Bajos (co-lead facilitator)
Chima Izugbara
Osmo Kontula
Cath Mercer (co-lead facilitator)
Chelsea Morroni
Richard de Visser
Georgina Yar-Oduro

**Hackathon Participants** – individuals who took part in the 3-day in-person hackathon in Kenya in January 2020.

Michele Andrasik
Soazig Clifton
Jennifer Toller Erausquin
Amanda Gabster
Amanda Gesselman
Martina Morris
Rocio Murad
Peterrock Muruiki
Martha Nicholson
Wendy Norman
Kate O'Connell
Adesola Olumide
Lisa Atieno Omond
Nicole Prause
Christopher Sengoga
Chantal Smith
Ariane van der Straten
Aleksandra Stuholfer
Alice Welbourn

**Steering Committee Members**
Linda Gail-Bekker
Laura Lindberg
Pedro Nobre

Emma Slaymaker
Annette Sohn
Kaye Wellings

**Supplementary file 6: Survey Implementation considerations**

**Survey Implementation Considerations.**

Below are some implementation considerations when designing, implementing, and disseminating findings from sexual and reproductive health survey research.

1. **Mapping stakeholders and organize the survey.** Often universities organize sexual health research studies, helping to ensure data management and implementation. Potential organizations include non-profit organizations and others. A local PI and local partners should be involved early in the process, mapping key national policy issues related to sexual health.
2. **Stakeholder engagement.** Engaging local stakeholders is essential. The rationale for stakeholder engagement is that this facilitates the research, helps translate to policy, and build capacity. Stakeholder engagement needs to cut across all phases of survey development and implementation.
3. **Identifying interviewers and interviewer training.** Need to find good interviewers and build longer-term capacity for research. Both recruiting from important subgroups and also have key subgroups to share and sensitize interviewers should be considered. Ideally data collectors should be specifically recruited and trained for this.
4. **Survey administration.** Survey will focus on interviewer administered, with some sections completed by the participant. Be aware that some people may be less familiar with computers, internet, and mobile phones. Support for interviewers should be considered related to safety (location of the interview, timing of the interview, mentorship); taking into consideration the interviewer's personal experiences.
5. **Protecting participants.** Practices to increase the likelihood of sexual minorities disclosing same sex behaviors, given high levels of homoprejudice in many settings. Avoiding triggers and allow participants to reinforce their own agency. Ensure that responses are confidential and not shared with anyone.
6. **Data management.** A plan and personnel are needed to ensure appropriate data cleaning, coding, and analysis. Use a secure server and back up data. A manual should include FAQs and explain how to solve common problems.

7. **Ethical review.** All research studies must be approved by a local ethical review board, in addition to other ethical review boards when relevant. Key ethical issues include the following: informed consent, confidentiality agreement, support for those with disabilities, voluntary participation (including ability to skip items), standardized protocols, minimizing or excluding identifiers, ethical review, dealing with sexual violence, ethics training, special populations (e.g., adolescents, pregnant women, incarcerated individuals, and others outlied in the stakeholder section).
8. **Translation and language.** The survey should be translated into the local language (involving key stakeholders) and back-translated to English. Need to identify appropriate local language(s).
9. **Pre-testing the survey.** Field testing is an umbrella term and includes tool testing ('pre-testing') for comprehension, essential and generally involves having some users and researchers discuss. This could include cognitive interviewing, formal validation, or going through the survey.
10. **Local support for victims of violence.** Individuals who have been identified as victims of violence should be connected to local resources (counselors, hotlines, community centers). However, given that formalized resources for sexual violence are rare in many LMIC settings, this point requires further consideration.
11. **Software/hardware**. Open data kit is open access software for collecting, managing, and using data in resource-constrained settings.  More details on ODK here and Kobo Toolkit here. Advise double-data entry if it has to be paper based. Include notes and constraints. Whatever you choose, ensure it is pilot-tested in your own context
12. **Sample size calculation.** The sample size of the survey should be calculated with a focus on the main purpose of the survey. For a simple cross-sectional survey, there are many open access tools (for example, OpenEpi has open access calculator that can be used offline). where you will have a hard time recruiting enough people to make meaningful comparisons.
13. **Dissemination.** Need to consider how to disseminate the findings of the research study. This can help with translation to policy-makers. This could include creating open access documents, creating messages for participants, and publishing in open access journals. One article here
